# Potential impacts of Washington State’s wildfire worker protection rule on construction workers

**DOI:** 10.1101/2021.07.19.21260289

**Authors:** Christopher Zuidema, Elena Austin, Martin A. Cohen, Edward Kasner, Lilian Liu, Tania Busch Isaksen, Ken-Yu Lin, June Spector, Edmund Seto

**Affiliations:** Department of Environmental and Occupational Health Sciences, University of Washington, Seattle, WA; Department of Construction Management, University of Washington, Seattle, WA; Department of Medicine, University of Washington, Seattle, WA

**Keywords:** forest fires, PM_2.5_, respirator, wildland fire, wildfire smoke protection rule

## Abstract

Driven by climate change, wildfires are increasing in frequency, duration, and intensity across the Western United States. Outdoor workers are being exposed to increasing wildfire-related particulate matter and smoke. Recognizing this emerging risk, Washington adopted an emergency rule and is presently engaged in creating a permanent rule to protect outdoor workers from wildfire smoke exposure. While there are growing bodies of literature on the exposure to and health effects of wildfire smoke in the general public and wildland firefighters, there is a gap in knowledge about wildfire smoke exposure among outdoor workers generally, and construction workers specifically, a large category of outdoor workers in Washington totaling 200,000 people. In this study, several data sources were linked including state-collected employment data and national ambient air quality data to gain insight into the risk of PM_2.5_ exposure among construction workers and evaluate the impacts of different air quality thresholds that would have triggered a new Washington emergency wildfire smoke rule aimed at protecting workers from high PM_2.5_ exposure. Results indicate the number of poor air quality days has increased in August and September in recent years. Over the last decade these months with the greatest potential for particulate matter exposure coincided with an annual peak in construction employment that was typically 9.4 to 42.7% larger across Washington counties (one county was 75.8%). Lastly, the “encouraged” threshold of the Washington emergency rule (20.5 μg/m^3^) would have resulted in 5.5 times more days subject to the wildfire rule on average across all Washington counties compared to its “required” threshold (55.5 μg/m^3^), and in 2020 the rule could have created demand for 1.35 million N-95 filtering facepiece respirators among construction workers. These results have important implications for both employers and policy makers as rules are developed. The potential policy implications of wildfire smoke exposure, exposure control strategies, and data gaps that would improve understanding of construction worker exposure to wildfire smoke are also discussed.

## INTRODUCTION

Wildfires in the Western US have increased in frequency and are burning greater land areas for longer periods of time (Balmes 2018). This trend, exacerbated by climate change (Abatzoglou and Williams 2016), is increasing the lengths of wildfire seasons (Reisen et al. 2015). In Washington (WA), where the summer climate is dry – especially east of the Cascade Mountain Range where conditions are hot and arid – environmental conditions contribute to wildfire ignition and spread. Smoke and pollution from wildfires can travel large distances from burns, including to urban areas (Wotawa 2000; Stefanidou et al. 2008; Reisen et al. 2015; Balmes 2018). The 2018 and 2020 wildfire seasons were particularly active, gaining media attention (Fields and Baruchman 2018; The Seattle Times 2020), prompting coordinated alerts from regional air quality and health agencies (PSCAA et al. 2018), and leading to mitigation planning in urban centers (Contreras 2019).

The composition of wildfire smoke is related to the environmental characteristics of the landscape that is burning (e.g., temperature, humidity, windspeed, fuel/forest type) (Stefanidou et al. 2008; Reisen et al. 2015; Balmes 2018) and has the potential to evolve as residential and commercial structures fuel burns. Incomplete wildfire combustion yields a large range of solid, liquid and gaseous pollutants (Stefanidou et al. 2008), including a range of potentially harmful pollutants such as silica, carbon monoxide, carbon dioxide, oxides of nitrogen and sulfur, methane, acrolein, formaldehyde, dioxins (Stefanidou et al. 2008), and hydrocarbons, including volatile organic compounds, polyaromatics, aldehydes, and furans (Materna et al. 1992; Slaughter et al. 2004; Statheropoulos and Karma 2007; Reisen et al. 2015). These constituents, as individual components or in mixture, may help explain recent observational and toxicological studies indicating wildfire smoke is more toxic than ambient particulate matter (PM) pollution (Kim Yong Ho et al. 2018; Aguilera et al. 2021). Despite the increased risk of wildfire smoke however, PM_2.5_ is still considered the main wildfire pollutant affecting human health (Schwela et al. 1999; Reid et al. 2005).

Occupational standards for PM exposure are not commonly exceeded for people, unless they are directly in the vicinity of the fire (e.g., OSHA’s 5 mg/m^3^ 8-hr time-weighted average for respirable dust), but the US Environmental Protection Agency (EPA) National Ambient Air Quality Standards (NAAQS) standards (e.g., PM_2.5_ >35 μg/m^3^ for a 24-hr period) can be exceeded by 1.2 to 10 times during wildfire events (Liu et al. 2015). Despite the large gap in these regulatory standards, resulting from the fact the EPA and OSHA standards have different human health and environmental protection aims and populations of interest, there is a notable lack of guidance for employers and employees related to wildfire smoke exposure. In fact, California (CA) is currently the only US state that has adopted permanent rules to protect non-firefighting workers from wildfire smoke exposure, including requirements for hazard identification, communication, training, and control of wildfire smoke exposure above an hourly Air Quality Index (AQI) level of 151 (PM_2.5_ = 55.5 μg/m^3^) (CA 2019). While other Northwest states – Oregon (OR) and WA – are currently engaged in similar rulemaking, their details, are only currently emerging. The WA Department of Labor and Industries (L&I) has implemented an emergency occupational wildfire protection rule for 2021, effective through November 2021, with an “encouraged” threshold of 20.5 μg/m^3^ (equivalent to a Washington Air Quality Advisory (WAQA) = 101 and AQI = 69) and a “required” threshold of 55.5 μg/m^3^ (WAQA = 173; AQI = 151) while continuing to work on a permanent rule. The rule proposed by OR Occupational Safety and Health (OSHA), has thresholds of 35.5 μg/m^3^ (AQI = 101) and 55.5 μg/m^3^ (AQI = 151) for successively stronger requirements to protect workers from wildfire smoke exposure (OR OSHA 2020). It is important to note that the wildfire smoke protection rules in CA, OR, and WA do not require the identification of wildfire smoke or wildfire-related PM_2.5_ specifically, yet rely on general ambient PM_2.5_ concentrations, for example as reported by government regulatory agencies.

A range of deleterious health effects have been associated with exposure to wildfire smoke. The most consistent evidence shows relationships with respiratory morbidity, specifically asthma exacerbations and chronic obstructive pulmonary disease, and a growing body of evidence of respiratory infections and all-cause mortality (Reid et al. 2016; Doubleday et al. 2020). Other potential outcomes include irritant reactions, such as headache, conjunctivitis, nasopharyngitis, sinusitis, tracheitis, and acute bronchitis (Shusterman et al. 1993); decreased lung function (Slaughter et al. 2004); and cardiovascular effects (Liu et al. 2015). Individuals with pre-existing conditions, specifically respiratory and cardiovascular, have been observed at higher risk (Liu et al. 2015). While these health effects have been studied in the general population, outdoor workers such as those in construction industries, may be at increased risk because many spend a considerable amount of time outside (Schulte and Chun 2009) and have a higher level of physical exertion and subsequent respiration rate than the general public. A recent meta-analysis (Kondo et al. 2019), suggested that more research is needed on general population sub-groups to establish unique effects, which may also be relevant among typically “healthy workers.” Furthermore, commercial and residential construction in WA is increasing – for example, from 39,021 residential units in 2000, to 48,4240 in 2019 (with a decrease to 43,881 in 2020 likely due to the COVID-19 pandemic) (US Census Bureau 2020) – reflecting an expanding construction workforce and at risk population in WA.

Construction workers already face many occupational hazards and are consistently subject to some of the highest rates of occupational accident, injury, and death. In 2018, the latest year with complete data, over 11.18 million construction workers made up 7.18% of the national workforce, yet accounted for 20.2% of fatalities and 5.8% of non-fatal injuries and illnesses (CDC 2020). In addition to traditional hazards (e.g., falls, electrocution, hearing loss, musculoskeletal disorders, and respiratory diseases (CPWR 2018)), construction workers are now exposed to increased PM from wildfires from the ambient environment. In conjunction with a growing construction workforce, seasonal trends in that workforce coincide with the wildfire season (summer in the Pacific Northwest), adding to the overall health burden. Exposure to or the health effects from wildfire smoke have been described in the general public (Shusterman et al. 1993; Reisen et al. 2015; Liu et al. 2015), agricultural workers (Austin et al. 2021), and wildland firefighters (Shusterman et al. 1993; Slaughter et al. 2004; Reinhardt and Ottmar 2004; Stefanidou et al. 2008; Aisbett et al. 2012; Adetona et al. 2013; Wu et al. 2021). However, no studies we are aware of have examined potential exposure to wildfire pollution among construction workers, especially as it is defined with PM_2.5_ thresholds under new wildfire smoke worker protection rules. There is therefore an important gap in the published literature on the impacts of wildfire smoke on construction workers. The aims of this study were to: 1) characterize the temporal patterns of poor air quality and construction employment across WA counties, 2) estimate potential exposure to high ambient PM_2.5_ concentrations among WA construction workers, 3) discuss the potential implications for state-level worker protection rulemaking in Washington, and 4) identify data gaps that would improve our understanding of the health risks and exposure to ambient air/wildfire pollution among WA construction workers.

## METHODS

### Study Area

The present analysis included all counties for Washington State, but three counties are highlighted: King, Spokane, and Yakima. These counties are a sample of the geographic variability in WA, and include a large metropolitan area (Seattle), rural and agricultural communities, and biomes east and west of the Cascade Mountain Range. These were also counties that bore a greater wildfire-related health burden in 2020, compared to other WA counties (Liu et al. 2021).

### Data Sources

#### Employment Data

Monthly employment data were gathered from the Washington Employment Security Department (ESD) Quarterly Census of Employment and Wages (QCEW) (WA ESD 2020). These data are collected cooperatively by the ESD and the US Bureau of Labor Statistics and report employment and wage information, in industries covered by unemployment insurance, by industry and county. Data are collected from quarterly unemployment tax forms filed by employers. Industries are categorized following North American Industrial Classification System (NAICS) codes. Data are considered of “excellent” accuracy/reliability, with only occasional interruptions due to employers being reclassified into a different industry or moving counties. Available data from non-farm monthly employment from 2002-2020 related to construction industries, which followed the 2002 2- and 3-digit NAICS codes were used to show longer term trends for all of WA, by plotting monthly employment totals for “construction” (NAICS sector code 23), “construction of buildings” (NAICS subsector code 236), “heavy and civil engineering construction” (NAICS subsector code 237), and “specialty trade contractors” (NAICS subsector code 238). For further analysis at the county level, ESD data were restricted to the construction sector for the 10-year period 2011-2020.

At the State level, more detailed information on the types of construction, in the form of NAICS national industry (6-digit) codes, was available from ESD QCEW. These data were restricted to industries within the construction sector (NAICS code 23) in 2020, from which the number of construction workers potentially engaged in outdoor construction and who would therefore be considered exposed to wildfire smoke was evaluated.

#### PM_2.5_ and Air Quality Index Data

PM_2.5_ data for WA were collected from the US Environmental Protection Agency (EPA) Air Quality System (AQS) for 2011-2020 (US EPA 2020), including measurements from all Federal Reference Method (FRM), Federal Equivalent Method (FEM), and non-FRM/FEM monitors and calculated daily PM_2.5_ averages for each county. Some WA counties do not have any monitoring sites, therefore no PM_2.5_ data were available for this part of this analysis. We used the concentration thresholds defined in proposed and promulgated worker protection rules, as well as the NAAQS PM_2.5_ standard, to identify counties and days impacted by wildfire smoke as described in the Data Analysis section below.

County-level daily Air Quality Index (AQI) data based on PM_2.5_ were also obtained from the AQS (US EPA 2020). The AQI is the EPA’s summary measure and communication tool for air pollution and level of health concern; it is informed by 5 pollutants: ground-level ozone, particle pollution, carbon monoxide, sulfur dioxide, and nitrogen dioxide. AQI levels are defined according to thresholds for each pollutant; for PM_2.5_ the levels are: “good” (AQI = 0-50; PM_2.5_ = 0-12.0 μg/m^3^), “moderate” (AQI = 51-100; PM_2.5_ = 12.1-35.4 μg/m^3^), “unhealthy for sensitive groups” (AQI = 101-150; PM_2.5_ = 35.5-55.4 μg/m^3^), “unhealthy” (AQI = 151-200; PM_2.5_ = 55.5-150.4 μg/m^3^), “very unhealthy” (AQI = 201-300; PM_2.5_ = 150.5-250.4 μg/m^3^), and “hazardous” (AQI ≥ 301; PM_2.5_ ≥ 250.5 μg/m^3^). This analysis was restricted to days with AQI levels defined by PM_2.5_, the pollutant defining days subject to wildfire protection rules. The number of days per month for each county with poor AQI levels ranging from “moderate” to “hazardous” were tallied, then averaged by month over the 2011-2020 period to estimate the mean number of days per month with AQI levels worse than “good.”

### Data Analysis

The combined analysis of air quality and construction employment was focused on the 10 years from 2011 through 2020. This period of time started following the multi-year economic recession beginning in 2008 and included the COVID-19 pandemic in 2020. To represent the annual cyclical pattern of construction employment, monthly ESD counts of construction workers were averaged over the 2011-2020 period, then a percent change from the month with the lowest count of construction workers as the reference point (January for most counties) was calculated. With this procedure, the average monthly change in the WA construction workforce at the county level was estimated. This change in monthly construction employment was plotted with 1) boxplots of mean daily PM_2.5_ concentration for each month and 2) the mean number of days with AQI warnings over the 2011-2020 period.

Although the wildfire smoke protection rules in CA, OR, and WA are intended to protect workers from wildfire smoke, as worded, they are based on exceedances of specified thresholds for ambient PM_2.5_ concentrations. The thresholds for each state were considered in our analysis as follows. For each county the number of days that exceeded several PM_2.5_ thresholds including 20.5 µg/m^3^ (AQI = 69; WAQA = 101, the “encouraged” threshold in the WA emergency rule (WA L&I 2021)); 35 µg/m^3^ (EPA NAAQS (US EPA 2016), which is also close to the 35.5 µg/m^3^ (AQI = 101) threshold proposed in OR (OR OSHA 2020); and 55.5 µg/m^3^ (AQI = 151), the first action level of the CA rule (CA 2019) and the “required” threshold in the WA emergency rule were tallied. Additionally, the number of days per month that exceeded 1) 20.5 µg/m^3^ (AQI = 69) for the 2011-2020 period and 2) 55.5 µg/m^3^ (AQI = 151) for 2020 were also tallied. The CA and WA rules are triggered by outdoor work of duration greater than one hour, above a threshold based on AQI as defined as EPA’s NowCast (an average of current and past concentrations over the prior 12 hours). The intent of EPA’s NowCast is to provide current air quality information that better reflects 24-hour exposures, for which much of the epidemiologic evidence is based (US EPA 2021). Because this analysis was retrospective, and thus, 24-hour average exposures can be computed, daily averages were used, which sufficiently reflects the intent of current AQI. For each county the Pearson correlation between 1) percent construction workforce county-level daily PM_2.5_ concentration and 2) percent construction workforce and the mean number of days with AQI warnings worse than “moderate,” each at the monthly time scale was computed. Wildfire exposure rules are applicable for either PM_2.5_ concentrations or PM_2.5_ AQI values, therefore both were included both in this analysis.

A map displaying construction employment according to 2020 ESD data by county and overlaid the AQS monitoring locations to illustrate the relationship between construction worker population and the degree to which WA counties have air quality data was prepared.

State-level employment data classified by 6-digit NAICS codes, were restricted in further analysis to the NAICS 2-digit sector code 23, focusing on the numbers of construction workers within 3-digit NAICS subsector codes 236, 237, and 238. The potential for outdoor work among these 6-digit construction codes were then evaluated, which would lead to increased exposure to wildfire-related smoke and PM.

To estimate construction worker-days of exposure to wildfire smoke in WA, for each county the number of construction workers at the beginning of each month were tabulated then multiplied by the number of days where PM_2.5_ in the county exceeded each threshold for each month, and summed across WA counties.

All data analysis was performed in R version 4.0.3.

## RESULTS

### Employment

Trends in construction employment from 2002 through 2020 for Washington State are shown in Figure 1 and for each WA county in Figure S1. Though King County had the largest number of construction workers, most counties generally followed similar long- and short-term trends. The number of construction workers declined dramatically in the recession that began in 2008, and after reaching a minimum in 2011 steadily increased through 2019 until the spring of 2020 where a sharp decrease then increase reflected the economic impacts of the COVID-19 pandemic. For example, in King County, in the “construction” sector there was a pre-recession high in September 2007 of 74,800, a February 2011 minimum of 43,300, and a high of 76,800 construction workers in August 2019. By the end of 2020, construction employment had nearly recovered to pre-pandemic levels. A distinct annual cyclical pattern in employment occurred throughout the time period, with the number of construction workers lowest during the winter months (December-February) and highest during the summer months (July-September). The distribution of construction workers by county for WA in 2020 is shown in Figure 2 (with the locations of EPA AQS monitors). The counties with the greatest number of construction workers were Snohomish, King, and Pierce Counties (in Western Washington) and Spokane County in Eastern Washington.

**Figure 1.**
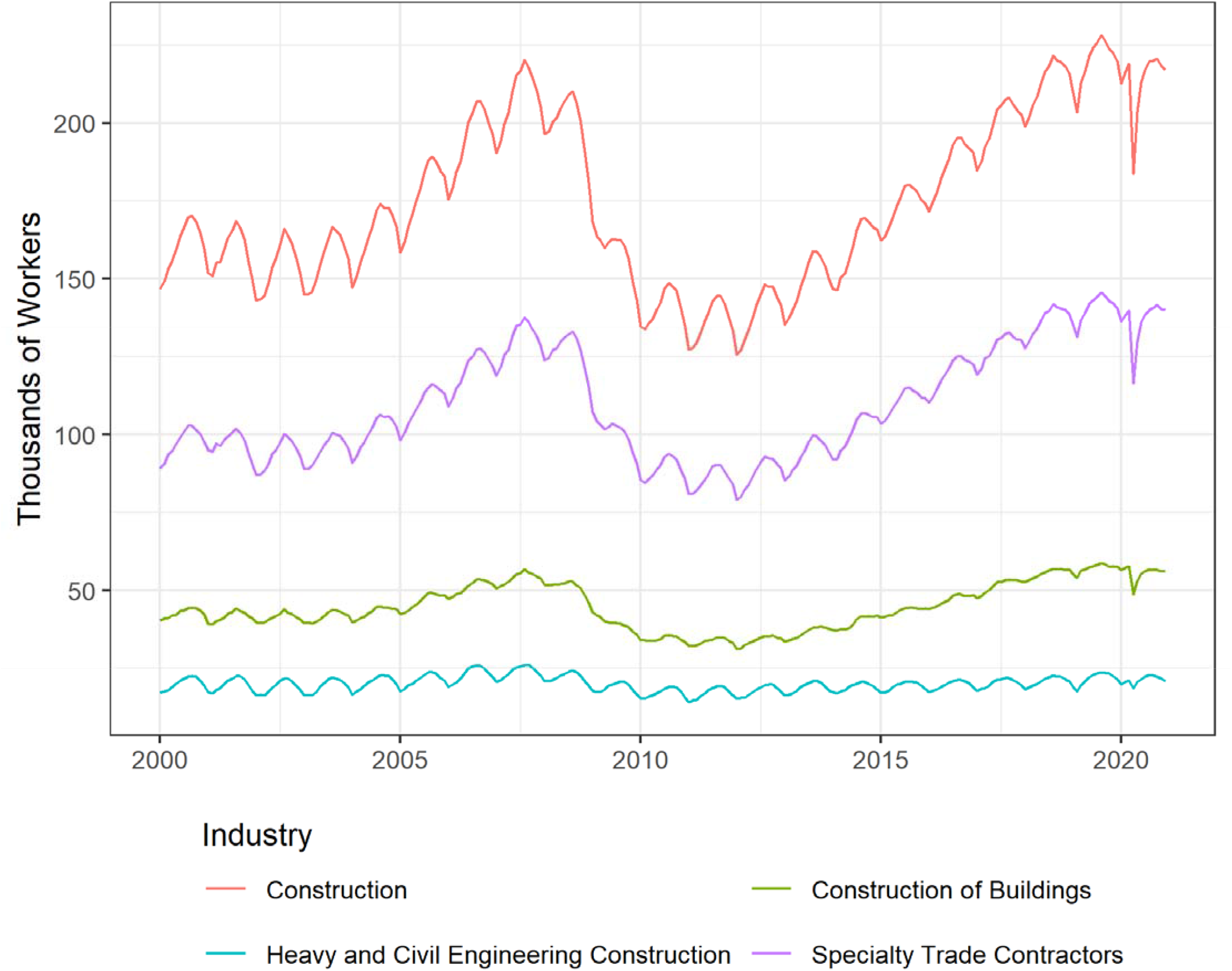
Monthly counts of Washington State construction workers. Construction of Buildings (NAICS code 236), Heavy and Civil Engineering Construction (NAICS code 237), and Specialty Trade Contractors (NAICS code 238) sum to Construction (NAICS code 23).

**Figure 2:**
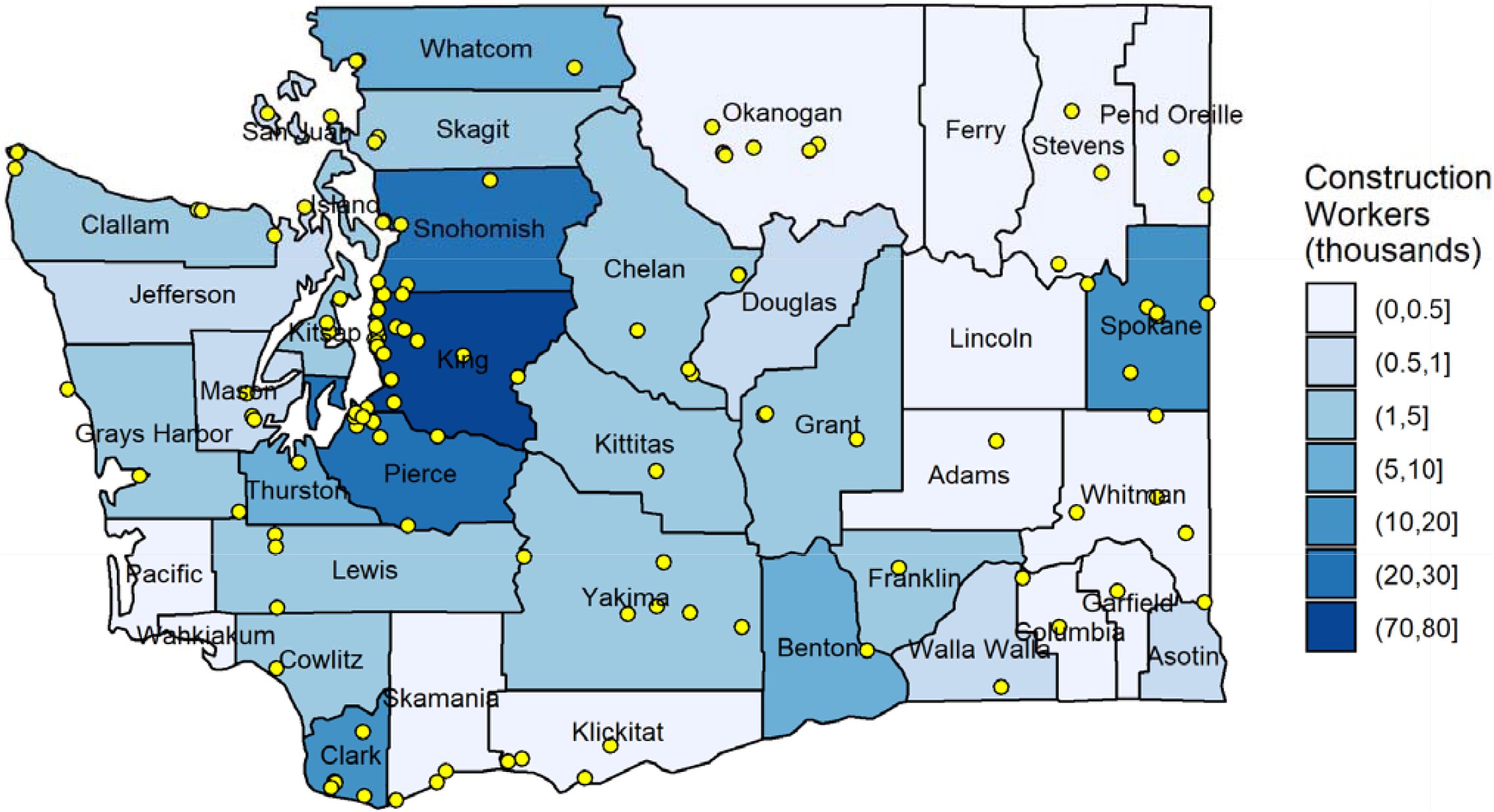
Map of Washington State with counties shaded according to construction employment (annual average of 2020 ESD data) and AQS monitor locations (points). (Note: construction employment for Garfield County was 2017 due to data availability.)

Tables 1 and S1 provide statewide detail about the 3- and 6-digit NAICS codes for construction workers in WA. Many construction workers have a high potential for outdoor work, and therefore exposure to ambient environmental conditions such as wildfire smoke, including civil and environmental engineering construction (NAICS code 237). Other types of construction, including construction of buildings (NAICS code 236) have a medium potential for outdoor work, which would largely depend on factors such as whether or not the heating, ventilation, and air conditioning (HVAC) system is operating and workers occupy indoor spaces supplied with filtered air. Specialty trade contractors (NAICS code 238) make up a large percent of WA construction workers, at 63.9% in 2020, and have mixed potential for outdoor work, largely depending on the trade. For instance, residential roofing contractors (NAICS code 238161) have a high potential for outdoor work, in contrast to residential finish carpentry contractors (NAICS code 238351). Collectively the NAICS codes assessed as having a high potential for outdoor work constitute 28.5% of construction workers in WA, while those with medium potential made up 68.1%.

**Table 1.** Summary of the number of construction workers by NAICS code in WA for 2020. (Note: table for all NAICS codes available in Supplementary Materials.)

| NAICS code | Industry | Potential<br>for<br>Outdoor<br>Work | Firms | Workers | Percent<br>of 2-digit<br>NAICS | Percent<br>of 3-digit<br>NAICS |
| --- | --- | --- | --- | --- | --- | --- |
| 23 | Construction |  | 26977 | 199784 | 100.0 |  |
| 236 | Construction of buildings |  | 9478 | 51636 | 25.8 | 100.0 |
| 236220 | Commercial building construction | Medium | 986 | 18808 | 9.4 | 36.4 |
| 236115 | New single family general contractors | Medium | 4338 | 14723 | 7.4 | 28.5 |
| 236118 | Residential remodelers | Medium | 3844 | 11908 | 6.0 | 23.1 |
|  | Other (NAICS 236116, 236117, 236210) | Medium | 310 | 6198 | 3.0 | 12.0 |
| 237 | Heavy and civil engineering construction |  | 1084 | 20576 | 10.3 | 100.0 |
| 237310 | Highway, street, and bridge construction | High | 239 | 6550 | 3.3 | 31.8 |
| 237110 | Water and sewer system construction | High | 320 | 4205 | 2.1 | 20.4 |
| 237130 | Power and communication system construction | High | 197 | 4195 | 2.1 | 20.4 |
|  | Other (NAICS 237120, 237210, 237990) | High | 329 | 5626 | 2.8 | 27.3 |
| 238 | Specialty trade contractors |  | 16416 | 127573 | 63.9 | 100.0 |
| 238212 | Nonresidential electrical contractors | Medium | 677 | 15418 | 7.7 | 12.1 |
| 238222 | Nonresidential plumbing and HVAC contractors | Medium | 460 | 14076 | 7.0 | 11.0 |
| 238221 | Residential plumbing and HVAC contractors | Medium | 1600 | 11992 | 6.0 | 9.4 |
|  | Other (NAICS codes 238211, 238311, 238321, 238911, 238912, 238161, 238312, 238111, 238351, 238131, 238992, 238292, 238991, 238322, 238162, 238112, 238171, 238122, 238331, 238152, 238341, 238142, 238392, 238352, 238141, 238151, 238192, 238391, 238132, 238332, 238121, 238291, 238191, 238172, 238342) | Mixed | 13684 | 86090 | 43.0 | 67.5 |

Over the 2011-2020 period, the construction workforce varied seasonally. For King, Spokane, and Yakima Counties, January was on average the month with the least number of construction workers (with the drop in April for King County resulting from inclusion of data during the 2020 COVID-19 pandemic). Using January as a baseline, the construction workforce increased throughout the year into summer where the workforce was an average of 9.4% larger in King County (September), and 23.7 and 26.2% larger in Spokane and Yakima Counties (August), respectively. For all counties in the State, the construction workforce was between 9.4 and 42.7% greater in summer, with Garfield County 75.8% larger.

### PM_2.5_ Air Pollution

PM_2.5_ varied over the course of the year for all WA counties (Figures 3 and S2). Among highlighted counties over the 2011-2020 period, the highest median daily PM_2.5_ concentrations were in Yakima County in winter (November = 12.0 µg/m^3^, December = 13.1 µg/m^3^, and January = 12.9 µg/m^3^). These winter concentrations were higher than the summer months of the wildfire season (July = 6.8 µg/m^3^, August = 8.5 µg/m^3^, and September = 7.5 µg/m^3^). A similar pattern existed for King and Spokane Counties, with median daily winter PM_2.5_ concentrations greater than summer, reflecting pollution from home heating (which in rural areas may be with wood-burning stoves or boilers), agricultural burning (as permitted by the State), and environmental conditions (e.g., atmospheric inversions). However, these elevated wintertime measures of central tendency, belie the more extreme daily concentrations observed, which occurred mostly in August and September. Over this 10-year period, months where the daily PM_2.5_ concentration exceeded the WA emergency wildfire rule’s “encouraged” threshold of 20.5 µg/m^3^ (AQI = 69) were generally August and September, but for some counties the rule may have also been applicable in months without wildfires (Table S2). Of the highlighted counties, Yakima exceeded 20.5 µg/m^3^, 46 and 50 days in August and September, respectively, over this 10-year period, compared to King County which experienced about one half to one third as many days. Of all WA counties, Okanagan had the greatest number of days (67 in August), followed by Chelan (59 in August) above 20.5 µg/m^3^ from 2011-2020. Similar results for 2020 are presented in Table S3 and incorporate annual estimates of construction employment with monthly results tabulated for the “required” threshold of 55.5 µg/m^3^ (AQI = 101). In 2020, most of the days that would have triggered this threshold in the WA rule were in September, corresponding with the major wildfire event.

**Figure 3.**
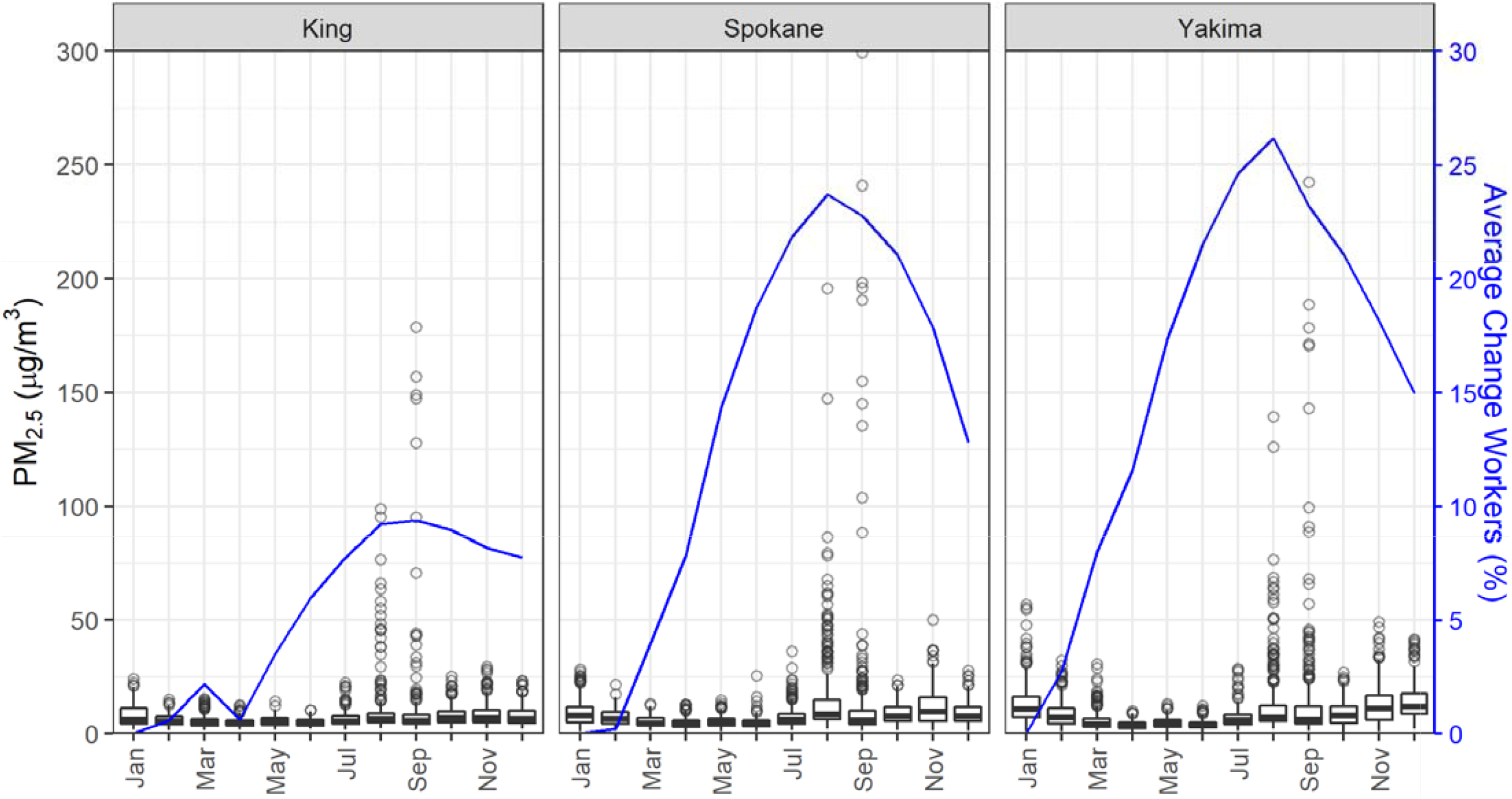
Daily PM_2.5_ concentrations and average monthly percent difference in construction workers from the month with the lowest number of workers for King, Spokane and Yakima, WA counties; 2011-2020. (Note: axes were restricted, omitting 2 and 3 data points above 300 µg/m^3^ for Spokane and Yakima Counties, respectively.)

### Air Quality Index

There was variability in the average number of days with AQI worse than “good” over the 2011-2020 period (Figures 4 and S3). Among highlighted counties, Yakima had the greatest number of poor air quality days, for all AQI categories “moderate” and worse (N = 1,196) and when restricting to the more severe AQI categories (i.e., omitting “moderate” AQI days; N = 147 days). For the “moderate” and worse days, this was 1.5 times greater than Spokane County and 1.8 times greater than King County, and for the more severe AQI days was 2.7 times greater for Spokane County and 3.7 times greater than King County. For all WA counties, all or nearly all of the days with the worst AQI levels (“very unhealthy,” “unhealthy,” and “hazardous”) occurred in August or September. There was also an increase in the number of days with poor AQI in more recent years (Figure S4), with the worst air quality days in August and September.

**Figure 4.**
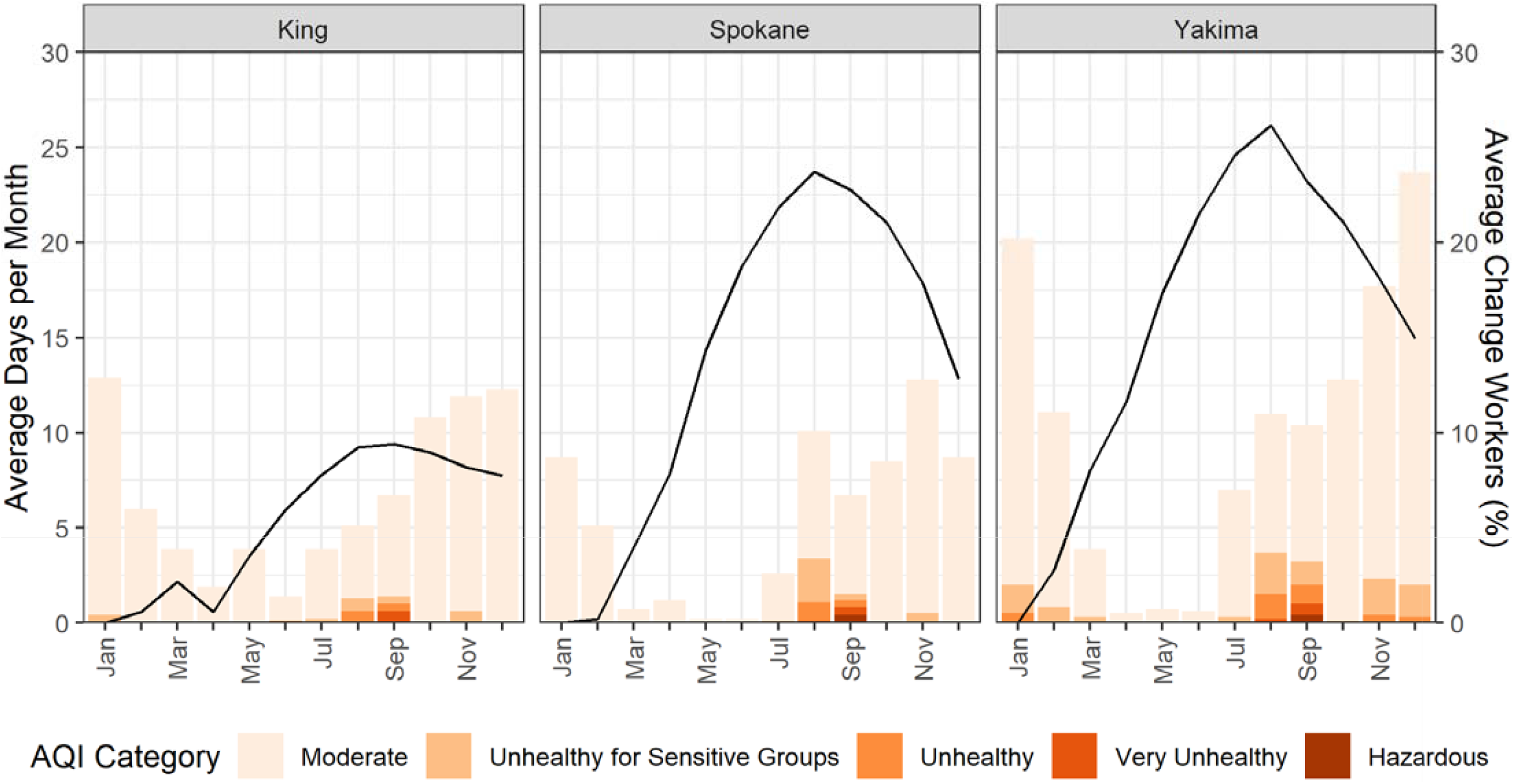
Average number of days per month with AQI worse than “good” and average monthly percent difference in construction workers from the month with the lowest number of workers for King, Spokane and Yakima, WA counties; 2011-2020.

### Relationship Between Air Quality and Seasonal Construction Workforce

Summaries of the relationship between seasonal construction employment and PM_2.5_ concentrations are presented in Figures 3 and S2. The months when the construction workforce is largest (August and September) coincide with months with the greatest number of high daily average PM_2.5_ concentrations. A similar pattern holds for construction employment and AQI (Figures 4 and Figure S3). Construction employment was generally highest in the summer months, when there were more days with higher AQI warnings.

Restricting AQI days to the most severe categories (“unhealthy for sensitive groups,” “unhealthy,” and “very unhealthy”), the Pearson correlation coefficients between the daily PM_2.5_ concentration or the average number of poor AQI days per month and the percent of the construction workforce is presented in Table S4. Among highlighted counties, there was moderately strong correlation between PM_2.5_ concentration and change in construction workforce for King County (p_King_ = 0.629), moderate correlation for Spokane County (p_Spokane_ = 0.501), but no correlation for Yakima County (p_Yakima_ = 0.109). Between the average number of days with AQI warnings and the change in construction workforce, the correlation was moderate for King and Spokane Counties (p_King_ = 0.517; p_Spokane_ = 0.508) but weak for Yakima County (p_Yakima_ = 0.196). Low to moderate correlations were observed for most WA counties, with the highest correlation between PM_2.5_ concentration and change in construction workforce (p_Garfield_ = 0.882) and between the average number of days with AQI warnings and the change in construction workforce (p_Garfield_ = 0.882). This observation with Garfield County however, may be influenced by the fact that there was no air quality data available prior to 2017 and recent years have been more impacted by wildfire smoke, and the large seasonal changes in the size of County’s construction workforce.

Estimated wildfire exposure, according to PM_2.5_ concentration thresholds, among WA construction workers is shown in Figure 5. Recent wildfire events in August 2017, August 2018, and September 2020 each resulted in more than 1 million construction worker-days of exposure for each of the three wildfire protection thresholds considered. As expected, the lowest threshold (20.5 µg/m^3^; AQI = 69) results in a larger number of worker-days of exposure, for example in August 2018, there were an estimated 2,330,000 construction worker-days of exposure compared to 880,500 construction worker-days under the 55.5 µg/m^3^ (AQI = 151) threshold. Additionally, the lower threshold also captured high pollution days in winter that were unlikely to be caused by wildfires. Extending the concept of construction worker-days of exposure to estimate the demand for respiratory protection in 2020 that could have been induced by the 55.5 µg/m^3^ (AQI = 69) threshold resulted in a calculated demand for filtering facepiece respirators totaling 1.35 million (Table S3) under the assumption that the annual average number of construction workers for each county would use one respirator for each day above the threshold.

**Figure 5.**
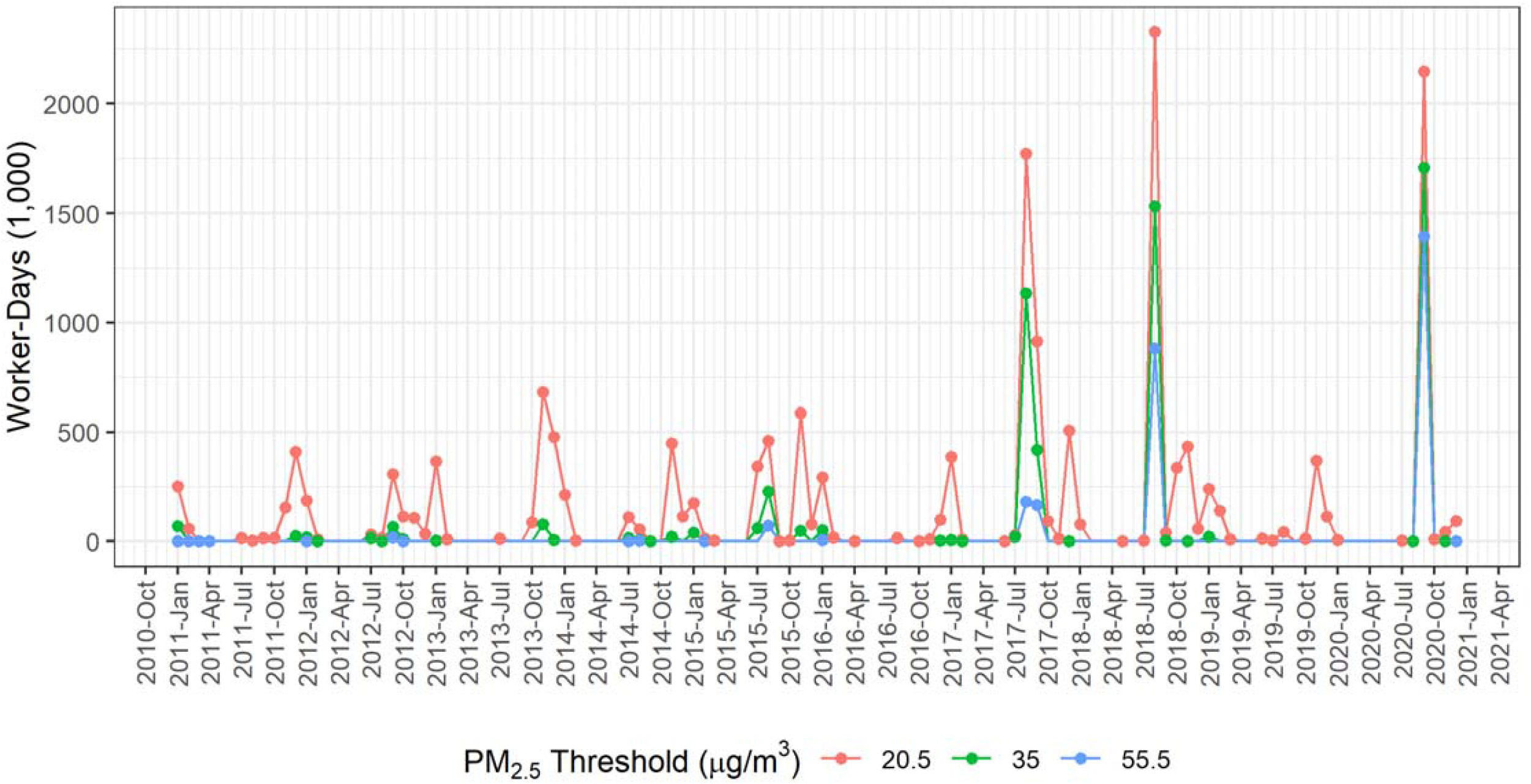
Estimated exposure to wildfire smoke among WA construction workers according to various PM_2.5_ thresholds.

## DISCUSSION

Even as PM_2.5_ concentrations have decreased across the US due to reduced industrial and vehicle emissions, the Northwest has not enjoyed the same improvements in air quality because of wildfires (Ford et al. 2018; McClure and Jaffe 2018). The continued influence of climate change is projected to increase wildfire-related PM_2.5_ as well as the associated effects on human health (Ford et al. 2018). As others have noted, workers such as agricultural and construction workers are at higher risk for wildfire smoke exposure, due to their prolonged outdoor work hours (Postma 2020; Austin et al. 2021). This analysis supports the conclusion that construction workers in WA face exposure to wildfire smoke. Furthermore, the cyclical nature of construction employment in WA means there are more construction workers on the job during summer months, at the same time when air quality is potentially poorer due to wildfire smoke and exposures are higher. With nearly 200,000 workers in WA employed in construction and construction-related industries in 2020 (WA ESD 2020), the impact is potentially quite large.

### Policy and Rulemaking

The status of outdoor workers in WA, including construction workers, came into clear focus in 2020 during the COVID-19 pandemic – many workers were classified as essential workers that continued to work outdoors during one of the State’s largest wildfire events in September. Additionally, since employers were not required to provide protection to workers exposed to wildfire smoke, either in the form of administrative or engineering controls or personal protective equipment (PPE), workers remained vulnerable to wildfire smoke inhalation. Against this backdrop, WA L&I had recognized that exposure to wildfire smoke posed a hazard to outdoor workers, specifically those in construction and agriculture, and is currently engaged in a permanent rule-making process aimed at protecting workers (296-62-085 WAC; General Occupational Health Standards) (WA L&I 2020). After the 2020 wildfire season, L&I fast-tracked an emergency rule for the 2021 wildfire season (enacted in mid-July 2021), as stakeholders debated a permanent rule based on a more or less stringent air quality standard (i.e., WAQA) compared to CA. CA’s worker protection rule for wildfire smoke is applicable when the PM_2.5_ AQI meets or exceeds 151 (PM_2.5_ ≥ 55.5 µg/m^3^), with directives for employers to provide and require employees to use N-95 respirators when AQI is above 500 (PM_2.5_ > 500.4 µg/m^3^) (CA 2019). The CA rule is based on current AQI as defined as EPA’s NowCast, which is an (unevenly) weighted average of current and past hourly concentrations over a 12-hour period and is used to assess “current” conditions until an entire day’s hourly concentrations have been monitored and the 24-hour average PM_2.5_ concentration can be used for the AQI calculation.

WA L&I’s emergency rule includes requirements for hazard communication of poor air quality levels and availability of protective measures; training, monitoring, and provisions for smoke-related health symptoms; and a hierarchy of controls that includes respiratory protection and engineering and administrative controls above a threshold of 55.5 µg/m^3^ (AQI = 151). At 20.5 µg/m^3^ (AQI = 69) however, employers are “encouraged” to implement exposure controls and are required to provide training. In this paper, these thresholds were examined retrospectively over the last 10 years, estimating the average number of days per month that would have triggered wildfire exposure protection requirements. In WA, September was the month most impacted by poor air quality due to wildfire smoke, followed by August – generally coinciding with peak construction workforces that are between 9.4 and 42.7% larger across WA counties.

Compared to CA and the “required” WA threshold, WA’s “encouraged” threshold of 20.5 µg/m^3^ (AQI = 69) is much lower, and if not clearly outlined when applicable, may trigger wildfire rule requirements during high air pollution events that are not related to wildfires. A limitation of the wording of the current WA rule is that high ambient PM_2.5_ concentrations above the specified thresholds that are from any source would technically trigger the rule’s requirements. For example, in many areas of WA, wood is used as a home heating fuel and the State permits agricultural burning; under certain atmospheric conditions common in winter months these practices may cause local concentrations to exceed 20.5 µg/m^3^ (AQI = 69). Our results demonstrate this potential, both in the daily average PM_2.5_ concentrations (Figures 3 and S2) and AQI (Figures 4 and S3) and in the weaker correlations between workers and PM_2.5_ and AQI for several counties. Among highlighted counties, Yakima is an example of a county with high PM_2.5_ concentrations unrelated to wildfires, and exemplifies the fact that PM_2.5_ doesn’t necessarily correspond to the summer months when there are high levels of construction activities and workers.

### Worker Protection and Controls

The WA emergency wildfire smoke protection rule draws on the hierarchy of controls to protect outdoor workers from wildfire smoke exposure. While encouraging employers to reduce employee exposure to wildfire smoke at PM_2.5_ concentrations above 20.5 µg/m^3^ (AQI = 69), employers must take action at 55.5 µg/m^3^ (AQI = 150) and work to reduce exposure below that level whenever feasible with engineering controls such as “enclosed buildings, structures, or vehicles where the air is adequately filtered.” When engineering controls insufficiently reduce employee exposure, administrative controls, such as work relocation, work schedule alterations, reduced work intensity, and increased rest periods should be implemented. Under the emergency rule, employers in WA would be encouraged to make respiratory protection (i.e., NIOSH-approved N-95 filtering facepiece respirators or KN-95 if N-95 is unavailable) available for voluntary use above 20.5 µg/m^3^ (AQI = 69), and would be required to above 55.5 µg/m^3^ (AQI = 150), avoiding requirements for fit testing and medical evaluation. For 2020 under the 55.5 µg/m^3^ (AQI = 150) threshold, this would have totaled 1.35 million respirators (Table S3). This estimate likely underestimates respirator demand because counties without PM_2.5_ concentration data (e.g., Douglas, Island, and Pacific, Counties among others) were not factored into this estimate, and overestimates respirator demand because some construction workers may be wearing respirators as normal practice, not all construction workers are outdoor workers (Tables 1 and S1), nor would all elect to wear a respirator for protection against wildfire exposure. The WA rule also requires improvements in medical surveillance and reporting of wildfire smoke in injury claims, which may help address current limitations in L&I data.

Beyond these measures, health and safety professionals can conduct research on and advocate for less traditional control strategies. For example, some construction workers are paid on a piece-rate basis, and a movement towards an hourly wage basis may reduce the physical exertion and corresponding exposure to wildfire smoke accompanying a faster work pace. In the agricultural sector there is some evidence that method of payment is associated with acute kidney injury (Moyce et al. 2017) and heat related illness (Spector et al. 2015), however this has not been studied in the construction industry for occupational wildfire smoke exposure. Employers also have an opportunity to combine training to related workplace hazards. For example, WA already has a rule protecting workers from outdoor heat (WAC 296-62-095), and because heat and wildfire exposure often coincide, employers could incorporate training about wildfire smoke and heat together. If there are diurnal patterns of air pollution, another administrative control strategy may be to pause work activities during parts of the day with higher concentrations. However, this requires employers to stay attuned to current local air quality conditions, rather than a daily AQI level or a forecasted AQI level for the next workday. Furthermore, workers without wage protection will still come to work in potentially unsafe conditions because they depend on the compensation. When it is not feasible to move all work indoors, another strategy may be to offer shelters or indoor spaces that are supplied with filtered air for rest periods and breaks, thus reducing exposure to wildfire smoke over the course of the day. Recent studies indicate that even consumer-grade portable air cleaners (i.e., non-commercial or non-industrial) can meaningfully reduce wildfire smoke concentrations indoors (Barn et al. 2008; Stauffer et al. 2020; Xiang et al. 2021).

### Study Limitations and Data Gaps

There were several limitations of this study, mostly related to limited data availability. Under the current occupational health paradigm, respirable PM fraction data for a large number of workers in different trades or industry categories at times with and without wildfire smoke exposure would ideally have been available. Worker exposure assessment to ambient air pollution may require a different approach, yet the complications of different PM size fractions (respirable PM with a 50% biologically based cut point of 4 µm versus PM_2.5_ with an instrument-based 2.5 µm cut point) and the continuous ambient and 8-hour occupational exposure workers face must be clearly addressed in future work. In this study, the EPA’s daily AQS PM_2.5_ and AQI data at the county level were used; these measurements were taken with the purpose of protecting public and environmental health, and result in a crude measure of exposure for WA construction workers. Even for this ecologic analysis there were data limitations, for example several WA counties did not have agency monitors for PM_2.5_. Future studies could leverage low-cost sensors to gather more extensive personal exposure data on wildfire and PM exposures as those tools develop (e.g., the US EPA and US Forest Service Fire and Smoke Map (US FS and US EPA 2020)).

Additional challenges related to data availability included the size of some counties (e.g., Garfield County), resulting in times within the study period without employment data. Similarly, several counties lacked air quality data (e.g., Douglas, Island, and Pacific Counties), which would require interpolation to estimate county-level air quality, or had incomplete data available (e.g., Garfield, Pend Oreille, and San Juan Counties) requiring averages that included periods without data. There was also a lack of health outcome data, preventing the study of health impacts of wildfire smoke exposure among WA construction workers.

As worker protection rules that focus on outdoor ambient conditions are promulgated (e.g., wildfire smoke and heat), there may be value in better characterizing numbers of outdoor workers for different NAICS codes and for other occupational classification systems, such as O*NET. In this study, for example, experience and judgement was used to evaluate the potential for outdoor work for construction NAICS codes. This qualitative approach avoided a number of further assumptions lacking supporting data that could be misleading, project a false sense of confidence, or induce errors in estimation, compared to a more quantitative model to estimate the proportion of outdoor workers. Potentially helpful pieces of information that would improve the characterization of outdoor workers is a standardized measure reporting whether or not a worker in particular occupation typically occupies a space supplied by filtered air or an HVAC system, or without a complete building envelope.

This study highlights the apparent conflict between occupational and environmental standards; whereas the general public may receive guidance on how to reduce exposures during wildfire smoke episodes there is a notable lack of work-specific guidance for employers and employees. Inconsistencies also exist among federal and state health-based guidance for ambient PM_2.5_, and states have implemented or proposed occupational thresholds derived from guidance that may be more stringent than federal standards. In WA for example, the WAQA index indicates that levels above 20.5 µg/m^3^ (AQI = 69) are unhealthy for sensitive groups, compared to the US EPA AQI that communicates similar risk for sensitive groups but at 35.5 µg/m^3^ (AQI = 101). Consistent guidance and messaging would help employers abide by occupational health requirements to protect their employees.

## CONCLUSION

Construction workers in Washington State are facing increased exposure to wildfires. Combined with long-term growth of the WA construction workforce, the annual cyclical nature results in a situation where more workers are exposed during the new “wildfire season” in August and September. This study retrospectively tallied the days that would have been subject to the WA L&I’s “encouraged” wildfire protection threshold of 20.5 μg/m^3^ (AQI = 69) over the last decade for each WA county and found it would result in 5.5 times more days subject to the wildfire protection rule than the WA “required” threshold of 55.5 μg/m^3^ (AQI = 151), especially if explicit provisions are not made to exclude high pollution days not associated with wildfires. The estimated demand for N-95 filtering facepiece respirators for construction workers in 2020 under WA’s emergency rule that could have been as high as 1.35 million. Our results can help inform both employers and policy makers as these rules are developed and, in some respects, can be generalized to other outdoor workers that the WA rule seeks to protect.

## Data Availability

All data in this study are publicly available secondary data accessible at the links below.

https://esd.wa.gov/labormarketinfo/covered-employment

https://esd.wa.gov/labormarketinfo/employment-estimates

https://aqs.epa.gov/aqsweb/airdata/download_files.html

## ACKNOWLEDGEMENTS

CZ was supported by the University of Washington’s Biostatistics, Epidemiology, and Bioinformatics Training in Environmental Health (BEBTEH), grant number T32ES015459, from the National Institute for Environmental Health Science (NIEHS). JTS was supported by CDC/NIOSH grant number T42OH008433.

## DATA AVAILABILITY STATEMENT

The data that support the findings of this study are available in the public domain: US EPA AQS (https://aqs.epa.gov/aqsweb/airdata/download_files.html) and WA ESD (https://esd.wa.gov/labormarketinfo/covered-employment and https://esd.wa.gov/labormarketinfo/employment-estimates).

## SUPPLEMENTAL MATERIALS

**Figure S1.**
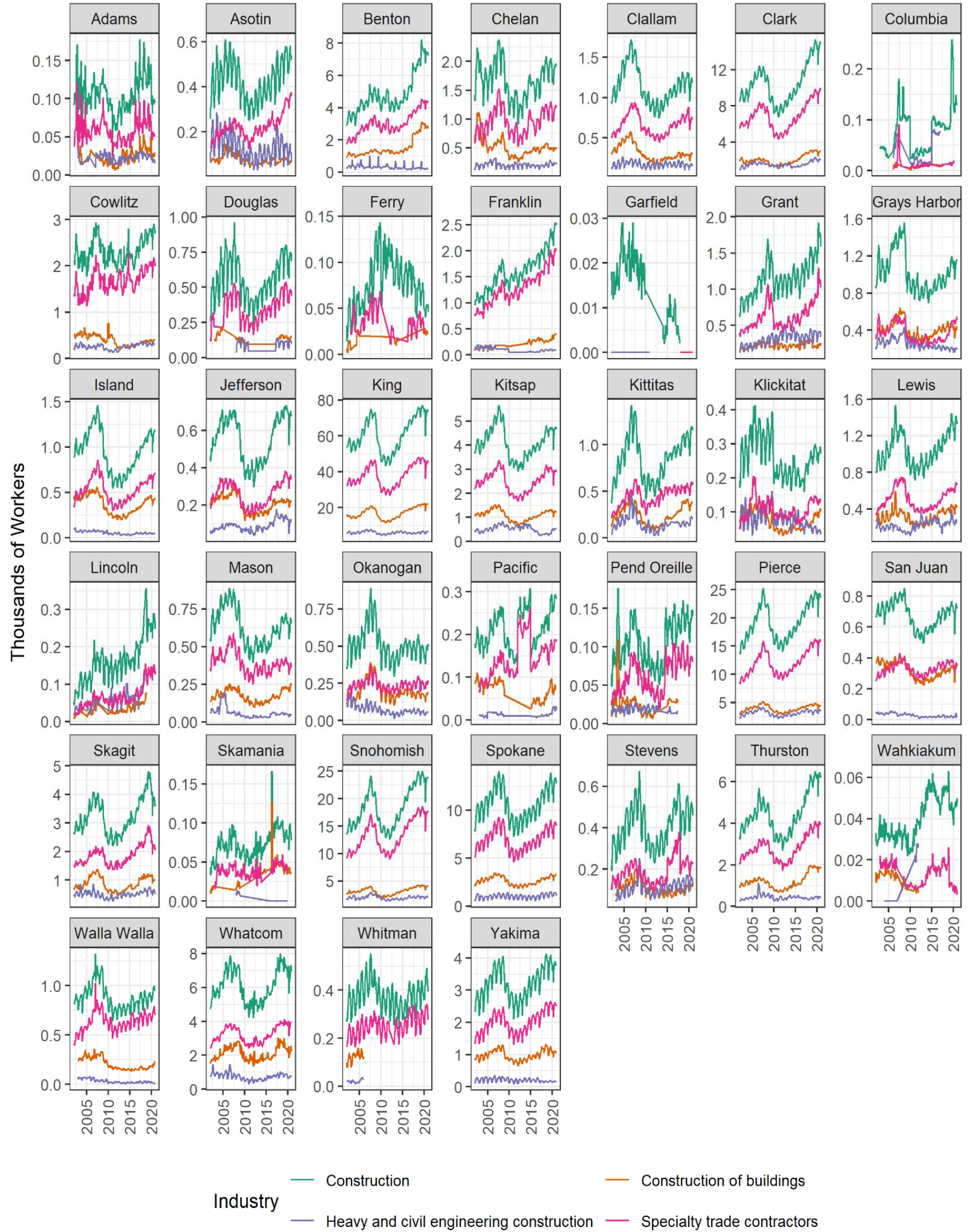
Monthly counts of WA construction workers.

**Figure S2.**
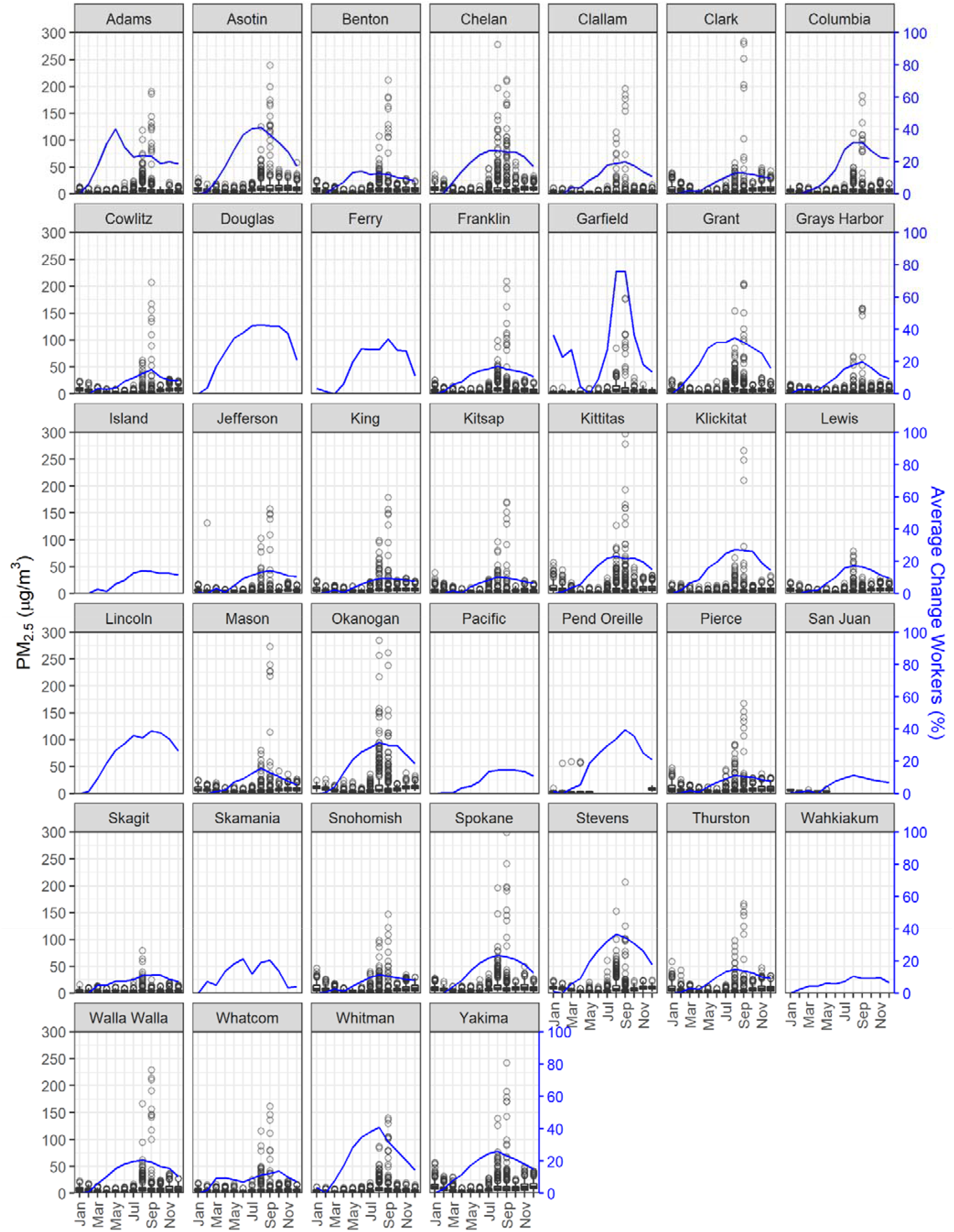
Mean daily PM_2.5_ concentration and average monthly percent difference in construction workers from the month with the lowest number of workers for all WA counties; 2011-2020. (Note: axes were restricted, omitting outlying data points above 300 µg/m^3^.

**Figure S3.**
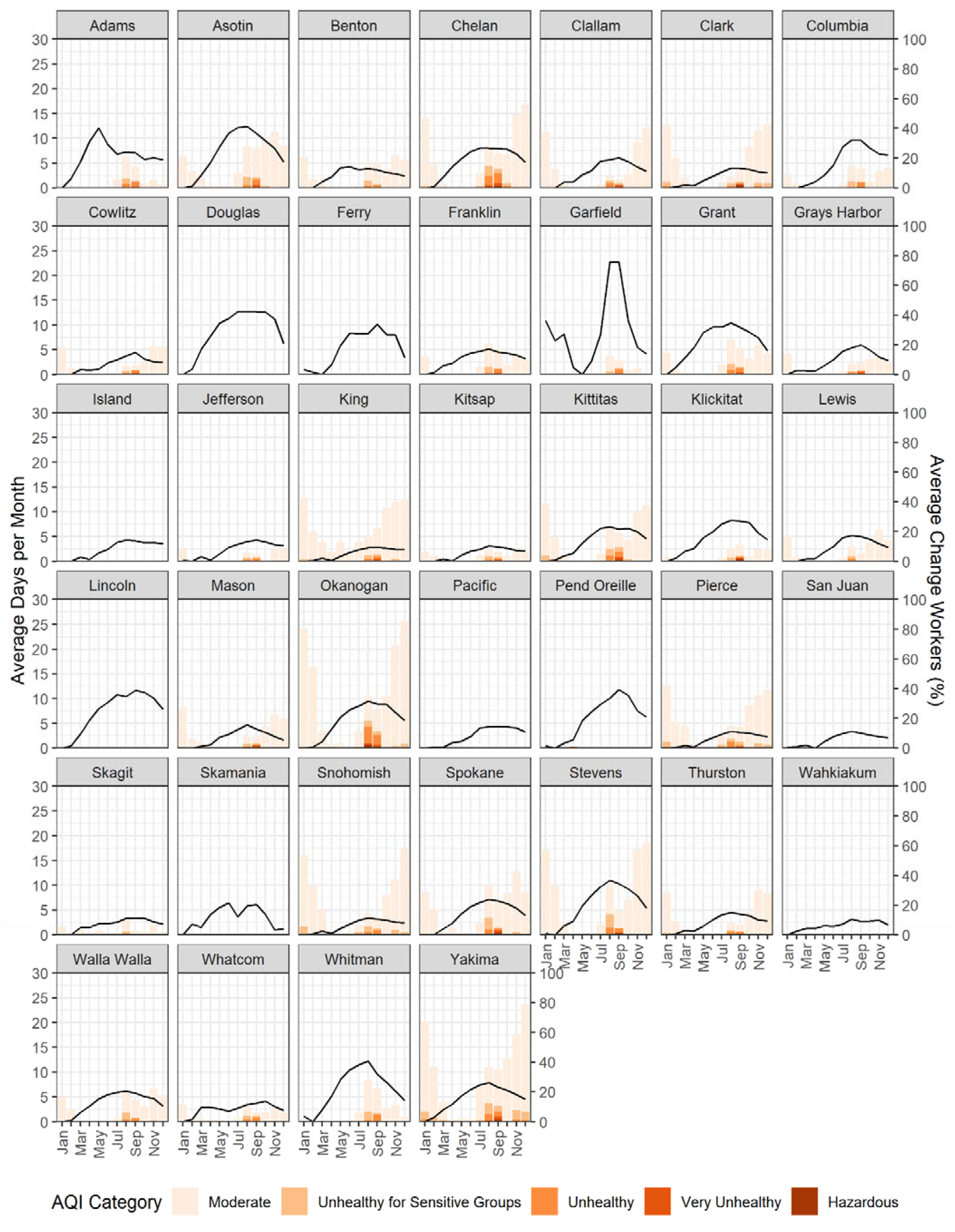
Average number of days per month with AQI worse than “good” and average monthly percent difference in construction workers from the month with the lowest number of workers averaged over 2011-2020 for all WA counties.

**Figure S4:**
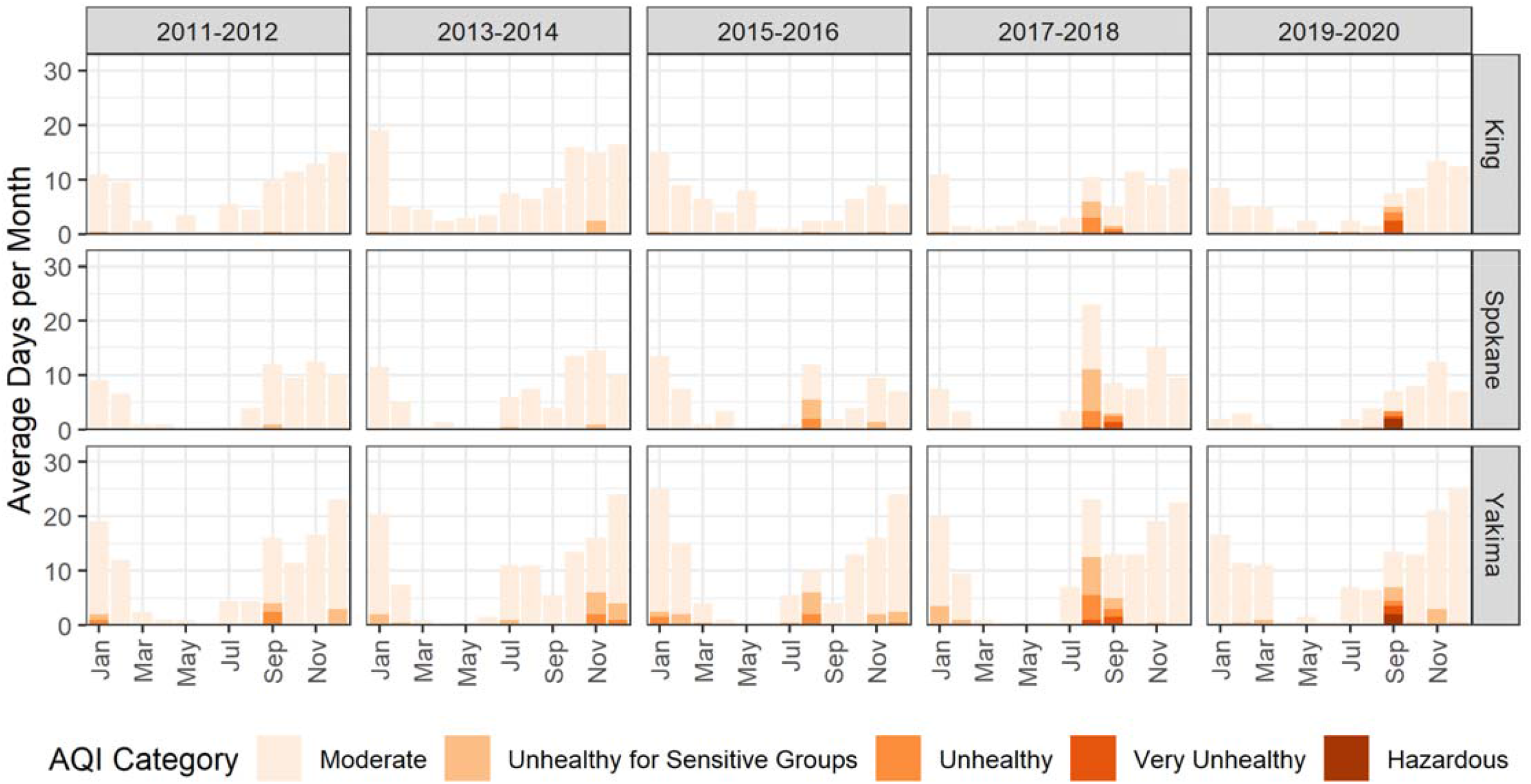
Average number of days per month with AQI warnings by county by month for 2-year periods; 2011-2020.

**Table S1.** Summary of the number of construction workers by NAICS code in WA for 2020.

| NAICS code | Industry | Potential<br>for<br>Outdoor<br>Work | Firms | Workers | Percent<br>of 2-digit<br>NAICS | Percent<br>of 3-digit<br>NAICS |
| --- | --- | --- | --- | --- | --- | --- |
| 23 | Construction |  | 26977 | 199784 | 100.0 |  |
| 236 | Construction of buildings |  | 9478 | 51636 | 25.8 | 100.0 |
| 236115 | New single family general contractors | Medium | 4338 | 14723 | 7.4 | 28.5 |
| 236116 | New multifamily general contractors | Medium | 58 | 1022 | 0.5 | 2.0 |
| 236117 | New housing for-sale builders | Medium | 192 | 1639 | 0.8 | 3.2 |
| 236118 | Residential remodelers | Medium | 3844 | 11908 | 6.0 | 23.1 |
| 236210 | Industrial building construction | Medium | 60 | 3537 | 1.8 | 6.8 |
| 236220 | Commercial building construction | Medium | 986 | 18808 | 9.4 | 36.4 |
| 237 | Heavy and civil engineering construction |  | 1084 | 20576 | 10.3 | 100.0 |
| 237110 | Water and sewer system construction | High | 320 | 4205 | 2.1 | 20.4 |
| 237120 | Oil and gas pipeline construction | High | 37 | 1169 | 0.6 | 5.7 |
| 237130 | Power and communication system construction | High | 197 | 4195 | 2.1 | 20.4 |
| 237210 | Land subdivision | High | 109 | 1117 | 0.6 | 5.4 |
| 237310 | Highway, street, and bridge construction | High | 239 | 6550 | 3.3 | 31.8 |
| 237990 | Other heavy construction | High | 183 | 3340 | 1.7 | 16.2 |
| 238 | Specialty trade contractors |  | 16416 | 127573 | 63.9 | 100.0 |
| 238111 | Residential poured foundation contractors | High | 905 | 4130 | 2.1 | 3.2 |
| 238112 | Nonresidential poured foundation contractors | High | 95 | 2230 | 1.1 | 1.7 |
| 238121 | Residential structural steel contractors | High | 30 | 458 | 0.2 | 0.4 |
| 238122 | Nonresidential structural steel contractors | High | 66 | 1923 | 1.0 | 1.5 |
| 238131 | Residential framing contractors | High | 748 | 3978 | 2.0 | 3.1 |
| 238132 | Nonresidential framing contractors | High | 78 | 645 | 0.3 | 0.5 |
| 238141 | Residential masonry contractors | High | 303 | 860 | 0.4 | 0.7 |
| 238142 | Nonresidential masonry contractors | High | 65 | 1077 | 0.5 | 0.8 |
| 238151 | Residential glass and glazing contractors | Medium | 142 | 847 | 0.4 | 0.7 |
| 238152 | Nonresidential glass and glazing contractors | Medium | 67 | 1403 | 0.7 | 1.1 |
| 238161 | Residential roofing contractors | High | 847 | 4749 | 2.4 | 3.7 |
| 238162 | Nonresidential roofing contractors | High | 93 | 2536 | 1.3 | 2.0 |
| 238171 | Residential siding contractors | High | 515 | 2111 | 1.1 | 1.7 |
| 238172 | Nonresidential siding contractors | High | 22 | 373 | 0.2 | 0.3 |
| 238191 | Other residential exterior contractors | High | 123 | 385 | 0.2 | 0.3 |
| 238192 | Other nonresidential exterior contractors | High | 85 | 786 | 0.4 | 0.6 |
| 238211 | Residential electrical contractors | Medium | 1470 | 7459 | 3.7 | 5.8 |
| 238212 | Nonresidential electrical contractors | Medium | 677 | 15418 | 7.7 | 12.1 |
| 238221 | Residential plumbing and HVAC contractors | Medium | 1600 | 11992 | 6.0 | 9.4 |
| 238222 | Nonresidential plumbing and HVAC contractors | Medium | 460 | 14076 | 7.0 | 11.0 |
| 238291 | Other residential equipment contractors | Medium | 61 | 417 | 0.2 | 0.3 |
| 238292 | Other nonresidential equipment contractors | Medium | 207 | 3231 | 1.6 | 2.5 |
| 238311 | Residential drywall contractors | Medium | 737 | 6533 | 3.3 | 5.1 |
| 238312 | Nonresidential drywall contractors | Medium | 115 | 4227 | 2.1 | 3.3 |
| 238321 | Residential painting contractors | Medium | 1846 | 6281 | 3.1 | 4.9 |
| 238322 | Nonresidential painting contractors | Medium | 194 | 2589 | 1.3 | 2.0 |
| 238331 | Residential flooring contractors | Medium | 890 | 1828 | 0.9 | 1.4 |
| 238332 | Nonresidential flooring contractors | Medium | 72 | 611 | 0.3 | 0.5 |
| 238341 | Residential tile and terrazzo contractors | Medium | 407 | 1368 | 0.7 | 1.1 |
| 238342 | Nonresidential tile and terrazzo contractors | Medium | 23 | 165 | 0.1 | 0.1 |
| 238351 | Residential finish carpentry contractors | Low | 894 | 4065 | 2.0 | 3.2 |
| 238352 | Nonresidential finish carpentry contractors | Low | 95 | 890 | 0.4 | 0.7 |
| 238391 | Other residential finishing contractors | Low | 93 | 691 | 0.3 | 0.5 |
| 238392 | Other nonresidential finishing contractors | Low | 139 | 997 | 0.5 | 0.8 |
| 238911 | Residential site preparation contractors | High | 1013 | 5172 | 2.6 | 4.1 |
| 238912 | Nonresidential site preparation contractors | High | 293 | 4777 | 2.4 | 3.7 |
| 238991 | All other residential trade contractors | Medium | 695 | 2976 | 1.5 | 2.3 |
| 238992 | All other nonresidential trade contractors | Medium | 256 | 3322 | 1.7 | 2.6 |

**Table S2.** Summary of the number of days that exceeded daily PM_2.5_ concentration thresholds for each WA county with PM_2.5_ data, 2011-2020.

| County | N >35 µg/m<sup>3</sup> | N >20.5 µg/m<sup>3</sup> | N >55.5 µg/m<sup>3</sup> | Days where PM<sub>2.5</sub> > 20.5 µg/m<sup>3</sup> |  |  |  |  |  |  |  |  |  |  |  |
| --- | --- | --- | --- | --- | --- | --- | --- | --- | --- | --- | --- | --- | --- | --- | --- |
|  |  |  |  | Jan | Feb | Mar | Apr | May | Jun | Jul | Aug | Sep | Oct | Nov | Dec |
| Adams | 34 | 73 | 21 | 0 | 0 | 0 | 0 | 0 | 1 | 2 | 46 | 23 | 0 | 1 | 0 |
| Asotin | 54 | 185 | 23 | 10 | 0 | 0 | 0 | 0 | 0 | 2 | 43 | 43 | 28 | 40 | 19 |
| Benton | 29 | 82 | 15 | 7 | 0 | 0 | 0 | 0 | 0 | 0 | 36 | 22 | 1 | 13 | 3 |
| Chelan | 76 | 171 | 45 | 8 | 2 | 0 | 0 | 0 | 0 | 6 | 59 | 45 | 11 | 23 | 17 |
| Clallam | 14 | 23 | 12 | 0 | 0 | 0 | 0 | 0 | 0 | 0 | 14 | 9 | 0 | 0 | 0 |
| Clark | 32 | 122 | 11 | 28 | 6 | 0 | 0 | 0 | 0 | 1 | 16 | 16 | 2 | 27 | 26 |
| Columbia | 25 | 47 | 12 | 0 | 0 | 0 | 0 | 0 | 0 | 0 | 25 | 15 | 0 | 7 | 0 |
| Cowlitz | 16 | 48 | 11 | 4 | 1 | 0 | 0 | 0 | 0 | 0 | 11 | 12 | 0 | 15 | 5 |
| Franklin | 29 | 71 | 14 | 2 | 0 | 0 | 0 | 0 | 0 | 1 | 37 | 22 | 0 | 7 | 2 |
| Garfield | 19 | 39 | 11 | 0 | 0 | 0 | 0 | 0 | 0 | 0 | 22 | 15 | 2 | 0 | 0 |
| Grant | 37 | 93 | 20 | 4 | 0 | 0 | 0 | 0 | 1 | 2 | 49 | 28 | 0 | 6 | 3 |
| Grays Harbor | 12 | 22 | 9 | 0 | 0 | 0 | 0 | 0 | 0 | 0 | 9 | 11 | 1 | 1 | 0 |
| Jefferson | 17 | 23 | 10 | 0 | 1 | 0 | 0 | 0 | 0 | 0 | 12 | 9 | 0 | 1 | 0 |
| King | 26 | 65 | 13 | 5 | 0 | 0 | 0 | 0 | 0 | 3 | 22 | 15 | 5 | 10 | 5 |
| Kitsap | 20 | 44 | 9 | 4 | 0 | 0 | 0 | 0 | 0 | 7 | 16 | 14 | 0 | 3 | 0 |
| Kittitas | 65 | 164 | 24 | 31 | 8 | 0 | 0 | 0 | 0 | 2 | 34 | 40 | 9 | 16 | 24 |
| Klickitat | 18 | 39 | 10 | 0 | 0 | 0 | 0 | 0 | 0 | 0 | 15 | 16 | 0 | 4 | 4 |
| Lewis | 9 | 36 | 5 | 1 | 0 | 0 | 0 | 0 | 0 | 0 | 18 | 3 | 1 | 11 | 2 |
| Mason | 19 | 64 | 10 | 10 | 0 | 0 | 0 | 0 | 0 | 0 | 17 | 15 | 3 | 12 | 7 |
| Okanogan | 76 | 155 | 47 | 6 | 1 | 0 | 1 | 1 | 0 | 8 | 67 | 40 | 0 | 12 | 19 |
| Pend Oreille | 5 | 5 | 5 | 0 | 1 | 1 | 3 | 0 | -- | -- | -- | -- | -- | -- | 0 |
| Pierce | 31 | 135 | 14 | 36 | 2 | 0 | 0 | 0 | 0 | 6 | 19 | 18 | 7 | 22 | 25 |
| San Juan | 0 | 0 | 0 | 0 | 0 | 0 | 0 | 0 | -- | -- | -- | -- | -- | -- | -- |
| Skagit | 5 | 14 | 3 | 0 | 0 | 0 | 0 | 0 | 0 | 1 | 12 | 1 | 0 | 0 | 0 |
| Snohomish | 28 | 140 | 10 | 33 | 5 | 0 | 0 | 0 | 0 | 5 | 22 | 15 | 0 | 29 | 31 |
| Spokane | 53 | 163 | 23 | 14 | 1 | 0 | 0 | 0 | 1 | 6 | 50 | 35 | 4 | 44 | 8 |
| Stevens | 53 | 107 | 22 | 4 | 0 | 0 | 0 | 0 | 0 | 4 | 57 | 23 | 1 | 16 | 2 |
| Thurston | 25 | 115 | 14 | 30 | 3 | 0 | 0 | 0 | 0 | 0 | 16 | 17 | 8 | 24 | 17 |
| Walla Walla | 33 | 80 | 15 | 3 | 0 | 0 | 0 | 0 | 0 | 0 | 34 | 19 | 6 | 12 | 6 |
| Whatcom | 22 | 40 | 10 | 0 | 0 | 0 | 0 | 0 | 0 | 2 | 22 | 13 | 0 | 3 | 0 |
| Whitman | 28 | 60 | 13 | 0 | 0 | 0 | 0 | 0 | 0 | 0 | 39 | 21 | 0 | 0 | 0 |
| Yakima | 70 | 279 | 27 | 47 | 16 | 4 | 0 | 0 | 0 | 6 | 46 | 50 | 12 | 49 | 49 |

**Table S3.**
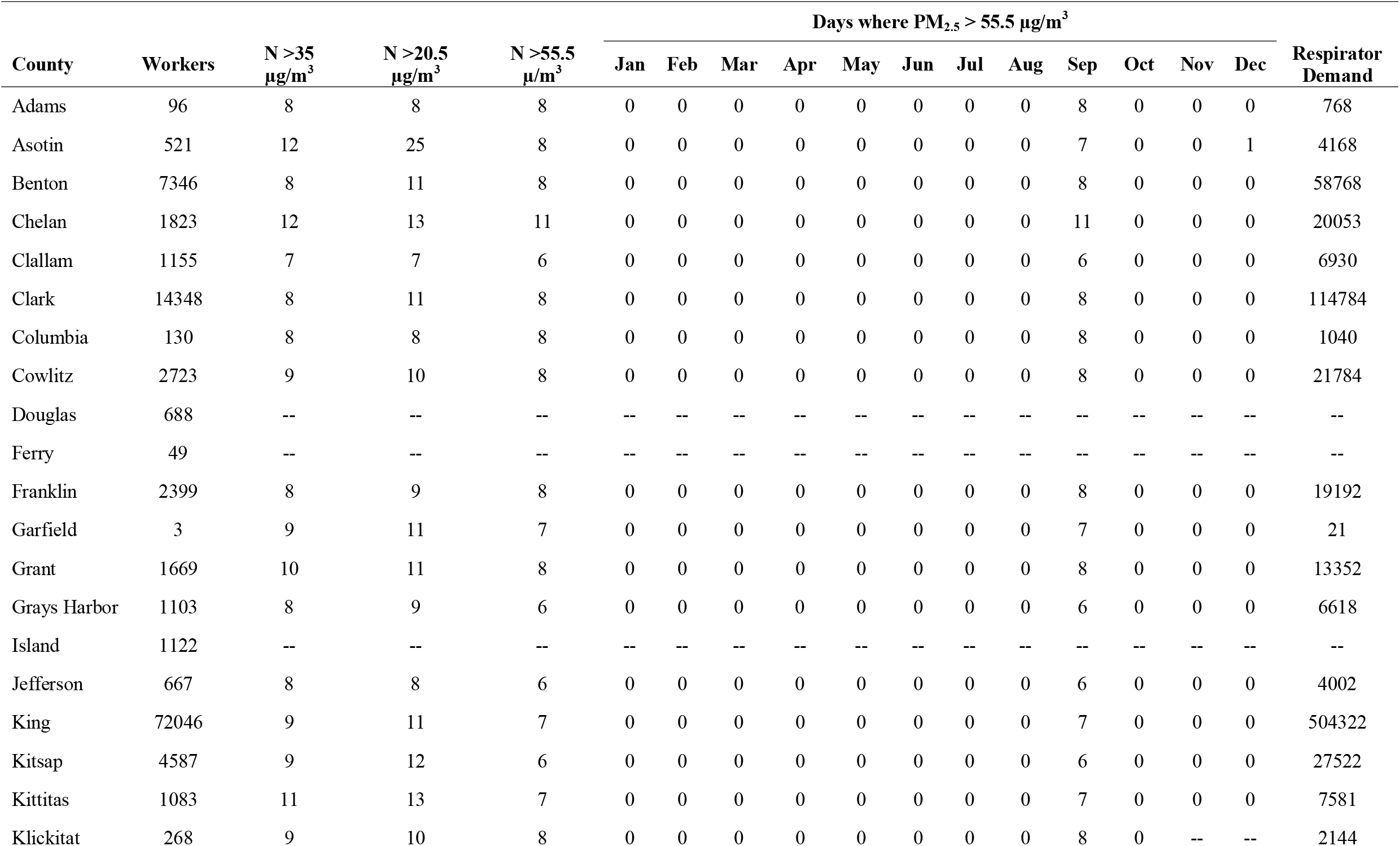
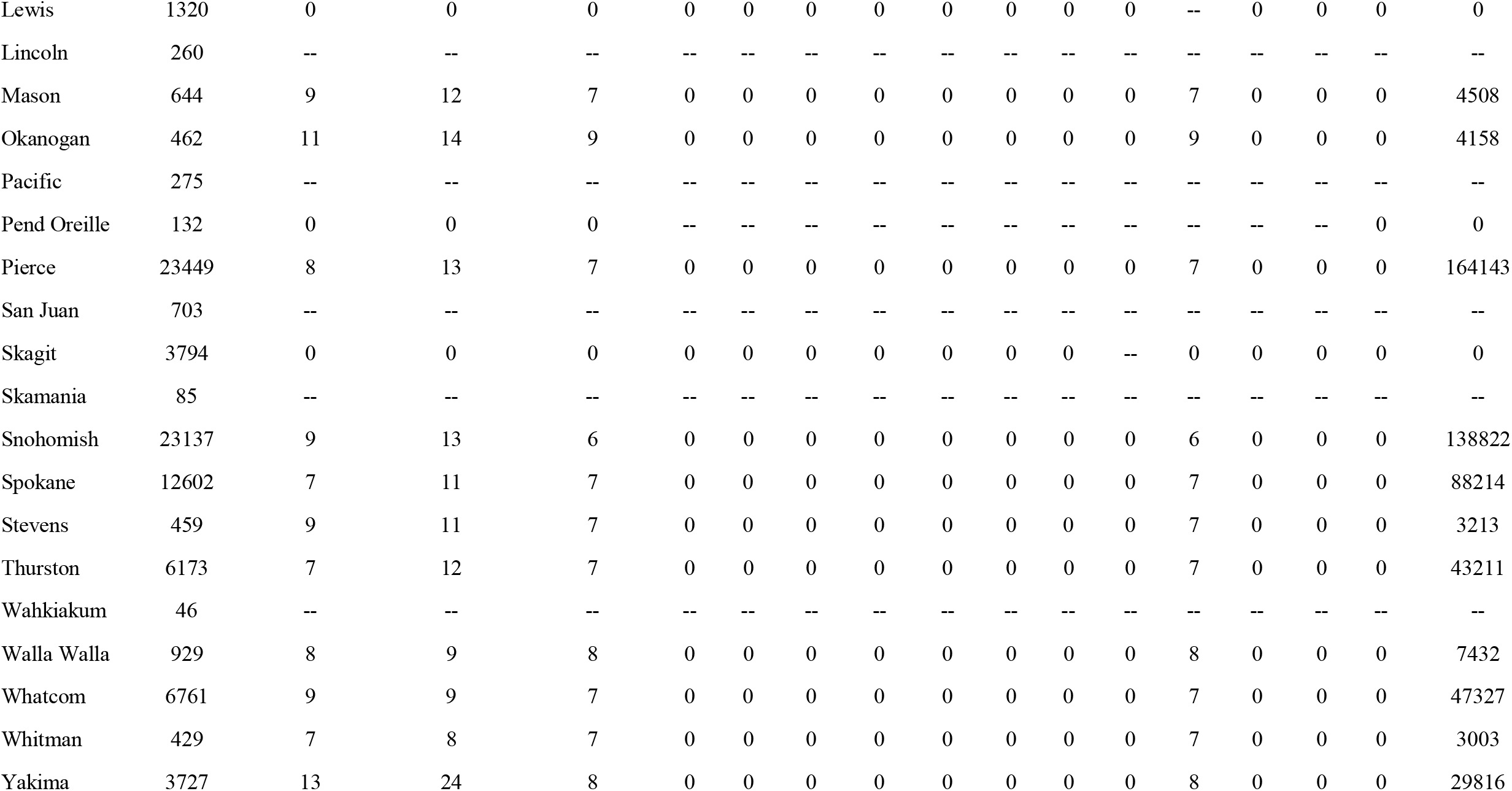
Summary of construction workers, the number of days per year that exceeded PM_2.5_ concentration thresholds, and the estimated demand for respiratory protection based on the 55.5 µg/m^3^ threshold for each WA county in 2020. (Note: construction employment for Garfield County was 2017 due to data availability.)

| County | Workers | N >35<br>µg/m<sup>3</sup> | N >20.5<br>µg/m<sup>3</sup> | N >55.5<br>µg/m<sup>3</sup> | Days where PM<sub>2.5</sub> > 55.5 µg/m<sup>3</sup> |  |  |  |  |  |  |  |  |  |  |  | Respirator<br>Demand |
| --- | --- | --- | --- | --- | --- | --- | --- | --- | --- | --- | --- | --- | --- | --- | --- | --- | --- |
|  |  |  |  |  | Jan | Feb | Mar | Apr | May | Jun | Jul | Aug | Sep | Oct | Nov | Dec |  |
| Adams | 96 | 8 | 8 | 8 | 0 | 0 | 0 | 0 | 0 | 0 | 0 | 0 | 8 | 0 | 0 | 0 | 768 |
| Asotin | 521 | 12 | 25 | 8 | 0 | 0 | 0 | 0 | 0 | 0 | 0 | 0 | 7 | 0 | 0 | 1 | 4168 |
| Benton | 7346 | 8 | 11 | 8 | 0 | 0 | 0 | 0 | 0 | 0 | 0 | 0 | 8 | 0 | 0 | 0 | 58768 |
| Chelan | 1823 | 12 | 13 | 11 | 0 | 0 | 0 | 0 | 0 | 0 | 0 | 0 | 11 | 0 | 0 | 0 | 20053 |
| Clallam | 1155 | 7 | 7 | 6 | 0 | 0 | 0 | 0 | 0 | 0 | 0 | 0 | 6 | 0 | 0 | 0 | 6930 |
| Clark | 14348 | 8 | 11 | 8 | 0 | 0 | 0 | 0 | 0 | 0 | 0 | 0 | 8 | 0 | 0 | 0 | 114784 |
| Columbia | 130 | 8 | 8 | 8 | 0 | 0 | 0 | 0 | 0 | 0 | 0 | 0 | 8 | 0 | 0 | 0 | 1040 |
| Cowlitz | 2723 | 9 | 10 | 8 | 0 | 0 | 0 | 0 | 0 | 0 | 0 | 0 | 8 | 0 | 0 | 0 | 21784 |
| Douglas | 688 | -- | -- | -- | -- | -- | -- | -- | -- | -- | -- | -- | -- | -- | -- | -- | -- |
| Ferry | 49 | -- | -- | -- | -- | -- | -- | -- | -- | -- | -- | -- | -- | -- | -- | -- | -- |
| Franklin | 2399 | 8 | 9 | 8 | 0 | 0 | 0 | 0 | 0 | 0 | 0 | 0 | 8 | 0 | 0 | 0 | 19192 |
| Garfield | 3 | 9 | 11 | 7 | 0 | 0 | 0 | 0 | 0 | 0 | 0 | 0 | 7 | 0 | 0 | 0 | 21 |
| Grant | 1669 | 10 | 11 | 8 | 0 | 0 | 0 | 0 | 0 | 0 | 0 | 0 | 8 | 0 | 0 | 0 | 13352 |
| Grays Harbor | 1103 | 8 | 9 | 6 | 0 | 0 | 0 | 0 | 0 | 0 | 0 | 0 | 6 | 0 | 0 | 0 | 6618 |
| Island | 1122 | -- | -- | -- | -- | -- | -- | -- | -- | -- | -- | -- | -- | -- | -- | -- | -- |
| Jefferson | 667 | 8 | 8 | 6 | 0 | 0 | 0 | 0 | 0 | 0 | 0 | 0 | 6 | 0 | 0 | 0 | 4002 |
| King | 72046 | 9 | 11 | 7 | 0 | 0 | 0 | 0 | 0 | 0 | 0 | 0 | 7 | 0 | 0 | 0 | 504322 |
| Kitsap | 4587 | 9 | 12 | 6 | 0 | 0 | 0 | 0 | 0 | 0 | 0 | 0 | 6 | 0 | 0 | 0 | 27522 |
| Kittitas | 1083 | 11 | 13 | 7 | 0 | 0 | 0 | 0 | 0 | 0 | 0 | 0 | 7 | 0 | 0 | 0 | 7581 |
| Klickitat | 268 | 9 | 10 | 8 | 0 | 0 | 0 | 0 | 0 | 0 | 0 | 0 | 8 | 0 | -- | -- | 2144 |
| Lewis | 1320 | 0 | 0 | 0 | 0 | 0 | 0 | 0 | 0 | 0 | 0 | 0 | -- | 0 | 0 | 0 | 0 |
| Lincoln | 260 | -- | -- | -- | -- | -- | -- | -- | -- | -- | -- | -- | -- | -- | -- | -- | -- |
| Mason | 644 | 9 | 12 | 7 | 0 | 0 | 0 | 0 | 0 | 0 | 0 | 0 | 7 | 0 | 0 | 0 | 4508 |
| Okanogan | 462 | 11 | 14 | 9 | 0 | 0 | 0 | 0 | 0 | 0 | 0 | 0 | 9 | 0 | 0 | 0 | 4158 |
| Pacific | 275 | -- | -- | -- | -- | -- | -- | -- | -- | -- | -- | -- | -- | -- | -- | -- | -- |
| Pend Oreille | 132 | 0 | 0 | 0 | -- | -- | -- | -- | -- | -- | -- | -- | -- | -- | -- | 0 | 0 |
| Pierce | 23449 | 8 | 13 | 7 | 0 | 0 | 0 | 0 | 0 | 0 | 0 | 0 | 7 | 0 | 0 | 0 | 164143 |
| San Juan | 703 | -- | -- | -- | -- | -- | -- | -- | -- | -- | -- | -- | -- | -- | -- | -- | -- |
| Skagit | 3794 | 0 | 0 | 0 | 0 | 0 | 0 | 0 | 0 | 0 | 0 | -- | 0 | 0 | 0 | 0 | 0 |
| Skamania | 85 | -- | -- | -- | -- | -- | -- | -- | -- | -- | -- | -- | -- | -- | -- | -- | -- |
| Snohomish | 23137 | 9 | 13 | 6 | 0 | 0 | 0 | 0 | 0 | 0 | 0 | 0 | 6 | 0 | 0 | 0 | 138822 |
| Spokane | 12602 | 7 | 11 | 7 | 0 | 0 | 0 | 0 | 0 | 0 | 0 | 0 | 7 | 0 | 0 | 0 | 88214 |
| Stevens | 459 | 9 | 11 | 7 | 0 | 0 | 0 | 0 | 0 | 0 | 0 | 0 | 7 | 0 | 0 | 0 | 3213 |
| Thurston | 6173 | 7 | 12 | 7 | 0 | 0 | 0 | 0 | 0 | 0 | 0 | 0 | 7 | 0 | 0 | 0 | 43211 |
| Wahkiakum | 46 | -- | -- | -- | -- | -- | -- | -- | -- | -- | -- | -- | -- | -- | -- | -- | -- |
| Walla Walla | 929 | 8 | 9 | 8 | 0 | 0 | 0 | 0 | 0 | 0 | 0 | 0 | 8 | 0 | 0 | 0 | 7432 |
| Whatcom | 6761 | 9 | 9 | 7 | 0 | 0 | 0 | 0 | 0 | 0 | 0 | 0 | 7 | 0 | 0 | 0 | 47327 |
| Whitman | 429 | 7 | 8 | 7 | 0 | 0 | 0 | 0 | 0 | 0 | 0 | 0 | 7 | 0 | 0 | 0 | 3003 |
| Yakima | 3727 | 13 | 24 | 8 | 0 | 0 | 0 | 0 | 0 | 0 | 0 | 0 | 8 | 0 | 0 | 0 | 29816 |

**Table S4.** Summary of Pearson correlation between the percent construction workforce and measures of air quality on the monthly timescale for Washington State counties, 2011-2020.

| County | Average AQI days per month | Average PM<sub>2.5</sub> (µg/m<sup>3</sup>) |
| --- | --- | --- |
| Adams | 0.119 | 0.04 |
| Asotin | 0.51 | 0.392 |
| Benton | 0.348 | 0.148 |
| Chelan | 0.469 | 0.215 |
| Clallam | 0.547 | 0.456 |
| Clark | 0.32 | 0.351 |
| Columbia | 0.59 | 0.637 |
| Cowlitz | 0.678 | 0.162 |
| Franklin | 0.465 | 0.428 |
| Garfield | 0.821 | 0.882 |
| Grant | 0.46 | 0.341 |
| Grays Harbor | 0.66 | 0.536 |
| Jefferson | 0.528 | 0.453 |
| King | 0.517 | 0.629 |
| Kitsap | 0.533 | 0.592 |
| Kittitas | 0.286 | 0.262 |
| Klickitat | 0.518 | 0.533 |
| Lewis | 0.443 | 0.267 |
| Mason | 0.658 | 0.115 |
| Okanogan | 0.467 | 0.194 |
| Pend Oreille | -0.401 | 0.246 |
| Pierce | 0.453 | 0.379 |
| San Juan | -- | -0.212 |
| Skagit | 0.333 | 0.459 |
| Snohomish | 0.334 | 0.106 |
| Spokane | 0.508 | 0.501 |
| Stevens | 0.506 | 0.232 |
| Thurston | 0.363 | 0.152 |
| Walla Walla | 0.462 | 0.245 |
| Whatcom | 0.404 | 0.277 |
| Whitman | 0.503 | 0.591 |
| Yakima | 0.196 | 0.109 |

## REFERENCES

Abatzoglou JT, Williams AP (2016) Impact of anthropogenic climate change on wildfire across western US forests. PNAS 113:11770–11775. https://doi.org/10.1073/pnas.1607171113

Adetona O, Simpson CD, Onstad G, Naeher LP (2013) Exposure of Wildland Firefighters to Carbon Monoxide, Fine Particles, and Levoglucosan. The Annals of Occupational Hygiene 57:979–991. https://doi.org/10.1093/annhyg/met024

Aguilera R, Corringham T, Gershunov A, Benmarhnia T (2021) Wildfire smoke impacts respiratory health more than fine particles from other sources: observational evidence from Southern California. Nat Commun 12:1493. https://doi.org/10.1038/s41467-021-21708-0

Aisbett B, Wolkow A, Sprajcer M, Ferguson SA (2012) “Awake, smoky, and hot”: Providing an evidence-base for managing the risks associated with occupational stressors encountered by wildland firefighters. Applied Ergonomics 43:916–925. https://doi.org/10.1016/j.apergo.2011.12.013

Austin E, Kasner E, Seto E, Spector J (2021) Combined Burden of Heat and Particulate Matter Air Quality in WA Agriculture. Journal of Agromedicine 26:18–27. https://doi.org/10.1080/1059924X.2020.1795032

Balmes JR (2018) Where There’s Wildfire, There’s Smoke. N Engl J Med 378:881–883. https://doi.org/10.1056/NEJMp1716846

Barn P, Larson T, Noullett M, Kennedy S, Copes R, Brauer M (2008) Infiltration of forest fire and residential wood smoke: an evaluation of air cleaner effectiveness. J Expo Sci Environ Epidemiol 18:503–511. https://doi.org/10.1038/sj.jes.7500640

CA (2019) California Code of Regulations, Title 8, Section 5141.1. Protection from Wildfire Smoke.

CDC (2020) CDC - Worker Health Surveillance - NIOSH Workplace Safety &Health Topics. https://www.cdc.gov/niosh/topics/surveillance/default.html. Accessed 29 Apr 2020

Contreras B (2019) Seattle prepares for health consequences of wildfire smoke. The Seattle Times

CPWR (2018) The Construction Chart Book. The Center for Construction Research and Training

Doubleday A, Schulte J, Sheppard L, Kadlec M, Dhammapala R, Fox J, Busch Isaksen T (2020) Mortality associated with wildfire smoke exposure in Washington state, 2006–2017: a case-crossover study. Environmental Health 19:4. https://doi.org/10.1186/s12940-020-0559-2

Fields A, Baruchman M (2018) Seattle pollution levels surge, as smoky air returns through at least Wednesday. The Seattle Times

Ford B, Martin MV, Zelasky SE, Fischer EV, Anenberg SC, Heald CL, Pierce JR (2018) Future Fire Impacts on Smoke Concentrations, Visibility, and Health in the Contiguous United States. GeoHealth 2:229–247. https://doi.org/10.1029/2018GH000144

Kim Yong Ho, Warren Sarah H., Krantz Q. Todd, King Charly, Jaskot Richard, Preston William T., George Barbara J., Hays Michael D., Landis Matthew S., Higuchi Mark, DeMarini David M., Gilmour M. Ian (2018) Mutagenicity and Lung Toxicity of Smoldering vs. Flaming Emissions from Various Biomass Fuels: Implications for Health Effects from Wildland Fires. Environmental Health Perspectives 126:017011. https://doi.org/10.1289/EHP2200

Kondo MC, De Roos AJ, White LS, Heilman WE, Mockrin MH, Gross-Davis CA, Burstyn I (2019) Meta-Analysis of Heterogeneity in the Effects of Wildfire Smoke Exposure on Respiratory Health in North America. International Journal of Environmental Research and Public Health 16:960. https://doi.org/10.3390/ijerph16060960

Liu JC, Pereira G, Uhl SA, Bravo MA, Bell ML (2015) A systematic review of the physical health impacts from non-occupational exposure to wildfire smoke. Environmental Research 136:120–132. https://doi.org/10.1016/j.envres.2014.10.015

Liu Y, Austin E, Xiang J, Gould T, Larson T, Seto E (2021) Health Impact Assessment of the 2020 Washington State Wildfire Smoke Episode: Excess Health Burden Attributable to Increased PM2.5 Exposures and Potential Exposure Reductions. GeoHealth 5:e2020GH000359. https://doi.org/10.1029/2020GH000359

Materna BL, Jones JR, Sutton PM, Rothman N, Harrison RJ (1992) Occupational Exposures in California Wildland Fire Fighting. American Industrial Hygiene Association Journal 53:69–76. https://doi.org/10.1080/15298669291359311

McClure CD, Jaffe DA (2018) US particulate matter air quality improves except in wildfire-prone areas. PNAS 115:7901–7906. https://doi.org/10.1073/pnas.1804353115

Moyce S, Mitchell D, Armitage T, Tancredi D, Joseph J, Schenker M (2017) Heat strain, volume depletion and kidney function in California agricultural workers. Occup Environ Med 74:402–409. https://doi.org/10.1136/oemed-2016-103848

OR OSHA (2020) Oregon Occupational Safety and HealthL: Rulemaking to Protect Employees from Unhealthy Levels of Wildfire SmokeL: Wildfire SmokeL: State of Oregon. https://osha.oregon.gov/rules/advisory/smoke/Pages/default.aspx. Accessed 13 Jul 2021

Postma J (2020) Protecting Outdoor Workers From Hazards Associated With Wildfire Smoke. Workplace Health Saf 68:52–52. https://doi.org/10.1177/2165079919888516

PSCAA, Kitsap Public Health District, Public Health Seattle &King County, Snohomish Health District, Tacoma-Pierce County Health Department (2018) Air Quality Alert for Puget Sound and Region Due to Wildfire Smoke

Reid CE, Brauer M, Johnston FH, Jerrett M, Balmes JR, Elliott CT (2016) Critical Review of Health Impacts of Wildfire Smoke Exposure. Environmental Health Perspectives 124:1334–1343. https://doi.org/10.1289/ehp.1409277

Reid JS, Koppmann R, Eck TF, Eleuterio DP (2005) A review of biomass burning emissions part II: intensive physical properties of biomass burning particles. Atmospheric Chemistry and Physics 5:799–825. https://doi.org/10.5194/acp-5-799-2005

Reinhardt TE, Ottmar RD (2004) Baseline Measurements of Smoke Exposure Among Wildland Firefighters. Journal of Occupational and Environmental Hygiene 1:593–606. https://doi.org/10.1080/15459620490490101

Reisen F, Duran SM, Flannigan M, Elliott C, Rideout K (2015) Wildfire smoke and public health risk. Int J Wildland Fire 24:1029. https://doi.org/10.1071/WF15034

Schulte PA, Chun H (2009) Climate Change and Occupational Safety and Health: Establishing a Preliminary Framework. Journal of Occupational and Environmental Hygiene 6:542–554. https://doi.org/10.1080/15459620903066008

Schwela DH, Goldammer JG, Morawska LH, Simpson O (1999) Health guidelines for vegetation fire eventsL: Guideline document. World Health Organization

Shusterman D, Kaplan JZ, Canabarro C (1993) Immediate health effects of an urban wildfire. West J Med 158:133–138

Slaughter JC, Koenig JQ, Reinhardt TE (2004) Association Between Lung Function and Exposure to Smoke Among Firefighters at Prescribed Burns. Journal of Occupational and Environmental Hygiene 1:45–49. https://doi.org/10.1080/15459620490264490

Spector JT, Krenz J, Blank KN (2015) Risk factors for heat-related illness in Washington crop workers. J Agromedicine 20:349–359. https://doi.org/10.1080/1059924X.2015.1047107

Statheropoulos M, Karma S (2007) Complexity and origin of the smoke components as measured near the flame-front of a real forest fire incident: A case study. Journal of Analytical and Applied Pyrolysis 78:430–437. https://doi.org/10.1016/j.jaap.2006.10.011

Stauffer DA, Autenrieth DA, Hart JF, Capoccia S (2020) Control of wildfire-sourced PM2.5 in an office setting using a commercially available portable air cleaner. Journal of Occupational and Environmental Hygiene 17:109–120. https://doi.org/10.1080/15459624.2020.1722314

Stefanidou M, Athanaselis S, Spiliopoulou C (2008) Health Impacts of Fire Smoke Inhalation. Inhalation Toxicology 20:761–766. https://doi.org/10.1080/08958370801975311

The Seattle Times (2020) Wildfire news updates, September 11: What to know today about the destructive fires in Washington state and on the West Coast. The Seattle Times

US Census Bureau (2020) US Census Bureau Building Permits Survey. https://www.census.gov/construction/bps/stateannual.html. Accessed 29 Apr 2020

US EPA (2020) The US Environmental Protection Agency, Technical Air Pollution Resources Air Quality System (AQS). https://aqs.epa.gov/aqsweb/airdata/download_files.html. Accessed 7 Nov 2020

US EPA (2016) National Ambient Air Quality Standards. https://www.epa.gov/criteria-air-pollutants/naaqs-table. Accessed 30 Jun 2021

US EPA (2021) AirNow - How is the NowCast algorithm used to report current air quality? https://usepa.servicenowservices.com/airnow?id=kb_article_view&sysparm_article=KB0011856. Accessed 13 Jul 2021

US FS, US EPA (2020) Fire and Smoke Map. https://fire.airnow.gov. Accessed 30 Jun 2021

WA ESD (2020) WA Employment Security Department. In: Covered Employment (QCEW). https://esd.wa.gov/labormarketinfo/covered-employment. Accessed 28 Apr 2020

WA L&I (2021) Wildfire Smoke Emergency Rule. https://www.lni.wa.gov/rulemaking-activity/AO21-26/2126CR103EAdoption.pdf. Accessed 16 Jul 2021

WA L&I (2020) Preproposal Statement of Inquiry. Washington State Department of Labor and Industries

Wotawa G (2000) The Influence of Canadian Forest Fires on Pollutant Concentrations in the United States. Science 288:324–328. https://doi.org/10.1126/science.288.5464.324

Wu C-M, Song C (Chuck), Chartier R, Kremer J, Naeher L, Adetona O (2021) Characterization of occupational smoke exposure among wildland firefighters in the midwestern United States. Environmental Research 193:110541. https://doi.org/10.1016/j.envres.2020.110541

Xiang J, Huang C-H, Shirai J, Liu Y, Carmona N, Zuidema C, Austin E, Gould T, Larson T, Seto E (2021) Field measurements of PM2.5 infiltration factor and portable air cleaner effectiveness during wildfire episodes in US residences. Science of The Total Environment 773:145642. https://doi.org/10.1016/j.scitotenv.2021.145642

